# Lifetime adversity exposure, mood symptoms, and immune mitochondrial bioenergetics

**DOI:** 10.64898/2026.06.02.26354718

**Authors:** Cynthia C. Liu, Catherine Kelly, Anna S. Monzel, Mandakh Bekhbat, Natalia Bobba-Alves, Veronica Ramirez, George M. Slavich, Robert-Paul Juster, Steve W. Cole, Martin Picard, Caroline Trumpff

## Abstract

Despite their prevalence, the pathophysiology of depression and anxiety remains poorly understood. Although adversity is a known risk factor, the mechanisms and biological contexts through which it contributes to mood disorder symptoms remain unclear. Immune and mitochondrial adaptations have both been implicated in mood disorders, suggesting the biological embedding of adversity may involve both systems. However, inconsistencies in the literature remain, partly due to reliance on mixed peripheral blood mononuclear cell (PBMC) populations despite substantial variability in mitochondrial biology across immune cell subtypes. We therefore investigated associations between adversity, mood disorder symptoms, immune cell proportions, and immune cell–specific mitochondrial bioenergetics (enzyme activities and respirometry) in participants from the Mitochondrial Stress, Brain Imaging, and Epigenetics (MiSBIE) study (n=105, age 18-60, 68% female, 35% with mitochondrial disease). Depressive and anxiety symptoms were positively associated with the monocyte-to-lymphocyte ratio, suggesting a shift toward greater innate relative to adaptive immunity. Associations between mood disorder symptoms and immune cell count were stronger in those exposed to greater early life adversity. Mood disorder symptoms were negatively associated with lymphocyte maximal mitochondrial respiratory capacity (MRC). As expected, the associations between mood disorder symptoms and lymphocyte mitochondrial bioenergetics (enzyme-based MRC and respiratory measurements) were stronger and more consistent among individuals exposed to higher lifetime adversity compared to those with lower lifetime adversity. Overall, these results suggest a complex interplay between adversity, immune cell mitochondrial bioenergetics, and mood disorder symptoms, highlighting immune mitochondrial biology as a potential allostatic pathway linking adversity to psychiatric disorders.

## 1. Introduction

Mood disorders, such as depression and anxiety, are among the most prevalent and persistent mental health conditions, representing a leading cause of disability worldwide and affecting a growing proportion of the population ^1,2^. These disorders have complex etiologies involving the biological embedding ^3^ of social-environmental factors, yet the mechanisms through which this embedding occurs remain unclear. Notably, stress ^4^ and adversity, particularly early-life adversity ^5–7^, have been shown to significantly increase the likelihood of developing depression and anxiety later in life ^8,9^. Although prior research has explored the impact of stress on mental health via biological systems, including the hypothalamic-pituitary-adrenal (HPA) axis ^10^ and inflammatory pathways ^7,11^, the molecular basis underlying these relationships remain partially unclear.

Stress and adversity have been linked to immune changes, yet findings are mixed. In individuals exposed to adversity, some studies report elevated total white blood cell (WBC) counts and granulocytes ^12^ along with decreased monocytes ^12^, while others find no significant differences as a function of stress ^13,14^. Stressor exposure has also been associated with increased innate immunity and decreased adaptive immunity ^15,16^, as well as with elevated inflammatory responses ^15,17–21^. These changes have, in turn, been observed in mood disorders. For instance, past studies have shown elevated WBC counts in depression ^22–24^, with specific increases in monocytes ^23,25–27^ and neutrophils ^22–24,27–29^, and decreases in lymphocytes ^22,26,27^.

Emerging evidence suggests that mitochondrial biology is also affected by stress and adversity. Studies in rodent models have demonstrated that stress exposure led to reduced mitochondrial oxidative phosphorylation (OxPhos) enzyme activity ^30–32^. In humans, stress and adversity have been associated with reduced brain mitochondrial enzymatic protein content ^33^. Stress and adversity have also been related to variation in blood mononuclear cell (PBMC) respiration, although directionality was mixed ^34–37^.

In the context of mood symptoms, patterns of reduced mitochondrial respiratory complex activity, gene expression, and enzymatic protein levels have been detected in major depressive disorder (MDD) and bipolar disorder in human post-mortem brain ^38–43^, muscle, and fibroblasts ^44–46^. In contrast, in immune cells, prior reports examining the association between immune cell mitochondrial biology and mood symptoms are mixed. Although some studies report no significant differences in PBMC OxPhos activity between healthy controls and patients with mood disorders ^47,48^, others report negative associations between depression severity and mitochondrial respiration in platelets ^49^, PBMCs ^50^, and T cells. Additionally, peripheral blood Complex I mRNA levels and platelet Complex IV activity have been reported to be respectively elevated in bipolar disorder ^51^ and positively associated with depressive symptoms ^49^.

Of relevance, exposure to stress and adversity may alter immune cell mitochondrial biology, thereby influencing the risk of developing mood disorders. Cross-species studies supporting this framework have identified overlapping mitochondrial gene expression changes in stress-exposed mice and humans with MDD ^52^. Human studies found interactions between childhood adversity and mitochondrial respiration in PBMCs of individuals with MDD ^35^, as well as between mood disorders, adversity and stress exposure and mitochondrial content in whole blood and PBMCs ^35,53^.

Despite advances in the literature, past research linking stress, adversity and mood disorders to immune cell mitochondrial biology is limited to mixed PBMCs. This is an important limitation because immune cell subtypes differ in their OxPhos profiles ^54–58^ and are differentially linked to stress ^59–61^ and mood disorders ^27^. Therefore, using mixed cell populations may confound variations in cell-type abundance with true mitochondrial alterations. Furthermore, whether life adversity influences the link between mood disorders and innate and adaptive cell immune proportions remains unclear.

To address these gaps, we examined associations between adversity, mood disorder symptoms, and circulating immune cell proportions and ratios of monocytes, neutrophils, platelets and lymphocytes. Next, we investigated whether adversity and mood symptoms were linked to mitochondrial bioenergetics in these same cell subtypes, via (1) maximal mitochondrial respiratory capacity (MRC), and (2) live mitochondrial respiration (Fig S1A). Lastly, we examined the influence of lifetime and childhood stressor exposure on the relation between mood symptoms and immune and mitochondrial parameters to investigate whether the development of adverse mood symptoms could be underpinned by imbalanced immune function and mitochondrial biology secondary to adversity exposure. We hypothesize that relationships between mitochondrial biology measures and mood symptoms may differ by adversity exposure.

## 2. Results

We assessed associations between immune cell type proportions, mitochondrial biology bioenergetics parameters, lifetime and childhood stressor exposure, and mood measures in a subset of healthy controls (n = 68) from the Mitochondrial Stress, Brain Imaging, and Epigenetics (MiSBIE) study ^62^ (Fig 1A), which we present in our main analyses. Additionally, these analyses were also conducted in the full MiSBIE study cohort (n = 105), which included patients with mitochondrial diseases (MitoD group, n = 37), offering a unique opportunity to examine whether these findings also apply in a larger sample with broader variation in mitochondrial biology. Given the exploratory nature of our study, we focus on unadjusted p-values in our analyses, however FDR-adjusted p-values (*p_adj_*) are also presented for reference. Demographic characteristics are presented in Table S1. Preliminary analyses showed that healthy controls did not differ from the MitoD group in demographics (i.e., age, sex, BMI, race/ethnicity), mood symptoms, or adversity measures.

**Figure 1.**
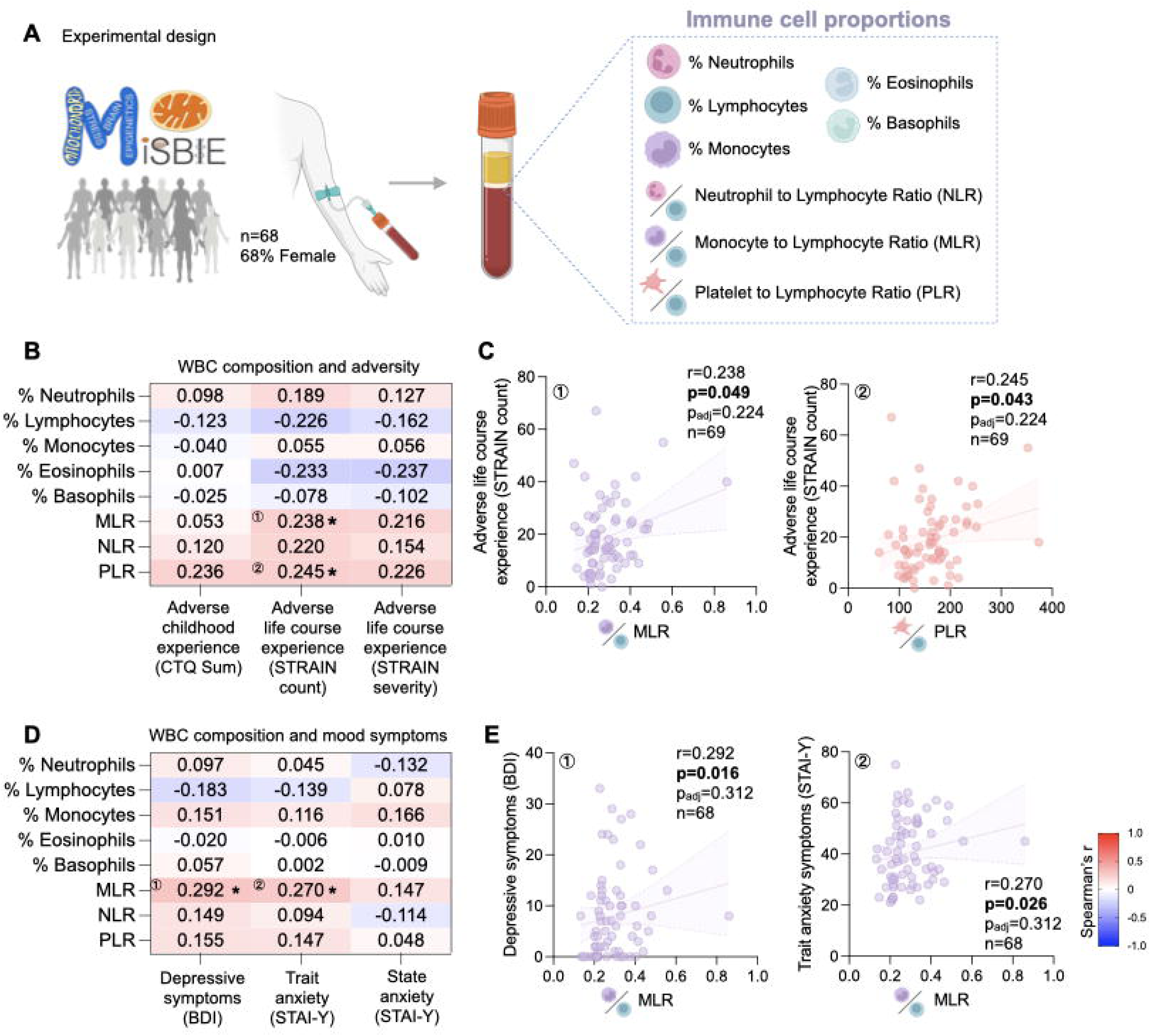
Associations between circulating white blood cell composition with mood symptoms and psychosocial adversity in controls. (A) Experimental design. Blood collected from MiSBIE control participants was used to quantify immune cell proportions. (B) Heatmap of Correlations between WBC percentages and ratios with measures of adversity (CTQ, STRAIN Total Count, STRAIN Total Severity). n=69. (C) Scatterplots showing correlations between WBC percentages and ratios with CTQ/STRAIN scores corresponding to the labeled comparisons on the heatmap. (D) Heatmap of correlations between white blood cell population (WBC) percentages and monocyte lymphocyte ratio (MLR), neutrophil lymphocyte ratio (NLR), platelet lymphocyte ratio (PLR) with measures of depression and anxiety symptoms (BDI, STAI-Trait, STAI - State). n=68. (E) Scatterplots showing correlations between WBC percentages and ratios with BDI/STAI scores corresponding to the labeled comparisons on the heatmap. In scatterplots, linear regression lines shown for illustrative purposes only, all correlations are Spearman rank correlations. Effect size and p-values from Spearman rank correlation. *p<0.05, **p<0.01, ***p<0.001, ****p<0.0001. No correlations remained significant following Benjamini-Hochberg FDR correction.

### Immune cell proportions and ratios and childhood/life adversity

The observed patterns of associations between immune cell type proportions and measures of adversity, while largely insignificant, suggest cell-type-specific associations with stressor exposures (Fig 1B-C, results for controls + MitoD are shown in Fig S1B). Both monocyte-to-lymphocyte ratio (MLR) (*r* = 0.24, *p* = 0.049, *p_adj_* = 0.22 Fig 1B-C) and platelet-to-lymphocyte ratio (PLR) (*r* = 0.25, *p* = 0.043, *p_adj_* = 0.22, Fig 1B, D) were positively correlated with lifetime stressor count as assessed by the Stress and Adversity Inventory (STRAIN), suggesting a shift in immune cell type proportion with adversity exposure.

### Immune cell proportions and ratios and depression/anxiety

The patterns of associations between immune cell type proportions and either symptoms of depression or state and trait anxiety were also cell-specific (Fig 1D-E, results including controls + MitoD are shown in S1C). As expected ^26,27^, positive associations were observed between MLR and depressive symptoms (*r* = 0.29, *p* = 0.016, *p_adj_* = 0.31). Additionally, MLR was also associated with trait anxiety symptoms (*r* = 0.27, *p* = 0.026, *p_adj_*= 0.31, Fig 1D-E). Both associations were also observed in the full study cohort including both healthy + MitoD participants (Fig S1C).

### Immune cell proportions and ratios and depression/anxiety, stratified by adversity

We further stratified participants into two groups based on their STRAIN lifetime severity scores and tested the interaction between immune cell proportions and depression/anxiety symptoms by STRAIN adversity group. Those with STRAIN severity scores above the median of all participants (> 44) were classified as the high lifetime adversity group and those with scores below the median (≤ 44) were classified as the low lifetime adversity group. We did not observe any significant moderating effects of lifetime adversity exposure on the relation between immune cell proportions and depression/anxiety symptoms in healthy controls (Table 1, Fig 2A-B). Results including both healthy controls and MitoD participants are shown in Fig S2A-B).

**Figure 2.**
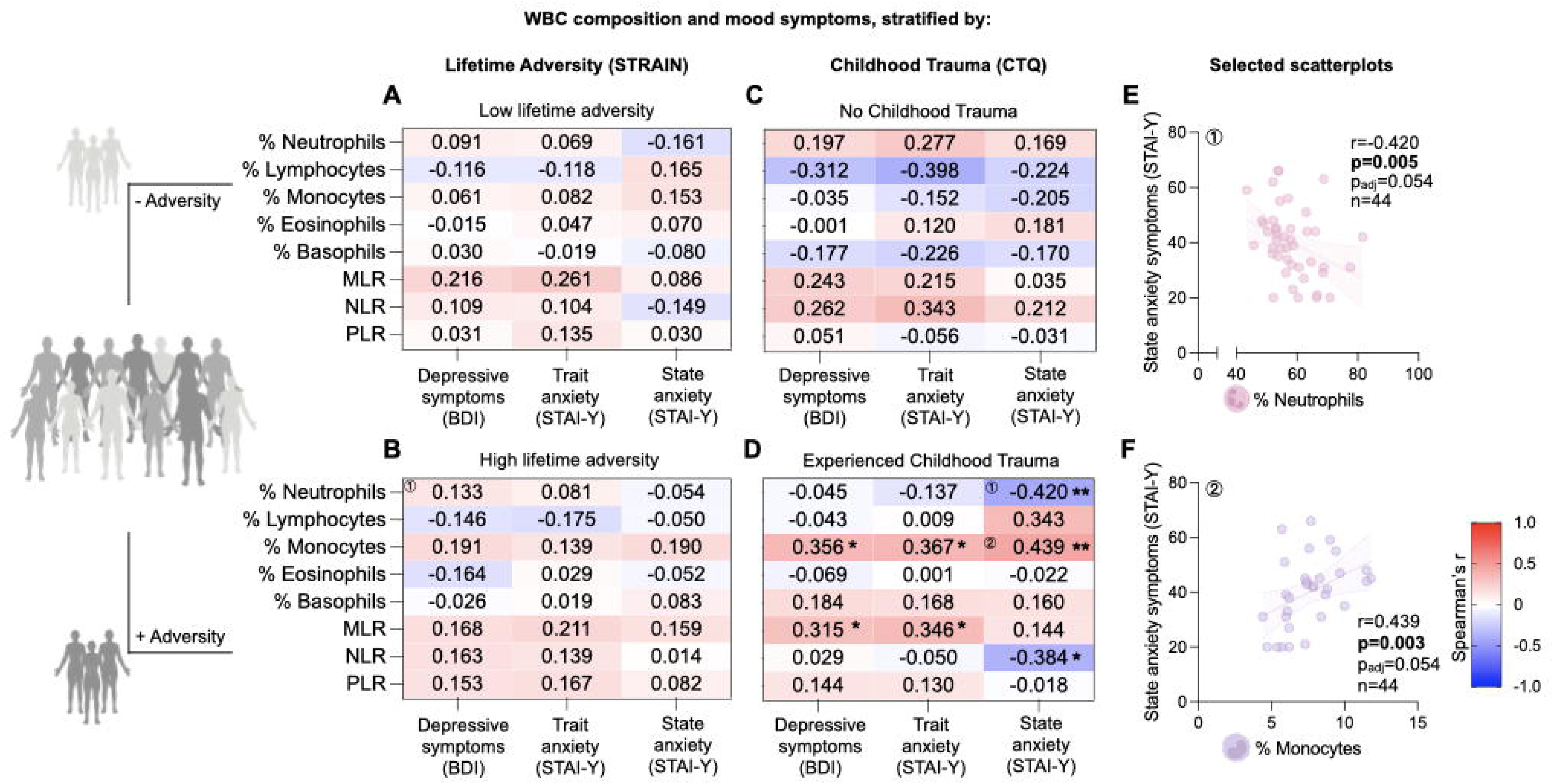
Associations between circulating white blood cell composition with mood symptoms and psychosocial adversity, stratified by adversity exposure. (A) Heatmap of correlations between white blood cell population (WBC) percentages and monocyte lymphocyte ratio (MLR), neutrophil lymphocyte ratio (NLR), platelet lymphocyte ratio (PLR) with measures of depression and anxiety symptoms(BDI, STAI-Trait, STAI - State) in individuals who experienced low lifetime adversity (STRAIN severity score ≤44). n=36-39. (B) Heatmap of correlations between WBC percentages and ratios with measures of depression and anxiety symptoms (BDI, STAI-Trait, STAI-State) in individuals who have experienced high lifetime adversity (STRAIN severity score >44). n=24=29. (C) Heatmap of correlations between WBC percentages and ratios with measures of depression and anxiety symptoms (BDI, STAI-Trait, STAI-State) in controls who did not experience childhood trauma. n=24. (D) Heatmap of correlations between WBC percentages and ratios with measures of depression and anxiety symptoms (BDI, STAI-Trait, STAI-State) in controls in individuals who experienced childhood trauma. n=44. (E) Scatterplot of correlation between NLR and state anxiety symptoms in controls in individuals who experienced childhood trauma. n=44. (F) Scatterplot of correlation between % Monocytes and state anxiety symptoms in controls in individuals who experienced childhood trauma. n=44. In scatterplots, linear regression lines shown for illustrative purposes only, all correlations are Spearman rank correlations. Effect size and p-values from Spearman rank correlation. *p<0.05, **p<0.01, ***p<0.001, ****p<0.0001. No correlations remained significant following Benjamini-Hochberg FDR correction.

**Table 1.** Effect of adversity exposure on the association between immune cell subtype proportion, MRC and Respirometry, and mood symptoms.

| Moderator: |  | Lifetime Adversity Exposure (STRAIN) |  |  |  | Childhood Adversity Exposure (CTQ) |  |  |  |
| --- | --- | --- | --- | --- | --- | --- | --- | --- | --- |
|  | Symptom | Interaction Coefficient Estimate | Interaction p-value | Interaction padj | n | Interaction Coefficient Estimate | Interaction p-value | Interaction padj | n |
| Immune Cell Proportions |  |  |  |  |  |  |  |  |  |
| Monocyte % | Depressive | 0.821 | 0.297 | 0.980 | 68 | 1.596 | 0.062 | 0.188 | 68 |
| Monocyte % | Trait anxiety | 0.98 | 0.449 | 0.980 | 68 | 2.466 | 0.053 | 0.188 | 68 |
| Monocyte % | State anxiety | 0.714 | 0.585 | 0.980 | 68 | 2.382 | 0.052 | 0.188 | 68 |
| Lymphocyte % | Depressive | 0.103 | 0.710 | 0.980 | 68 | 0.429 | 0.196 | 0.392 | 68 |
| Lymphocyte % | Trait anxiety | 0.011 | 0.980 | 0.980 | 68 | 0.721 | 0.147 | 0.320 | 68 |
| Lymphocyte % | State anxiety | -0.399 | 0.391 | 0.980 | 68 | **1.101** | **0.020** | 0.188 | 68 |
| Neutrophil % | Depressive | -0.144 | 0.537 | 0.980 | 68 | -0.425 | 0.127 | 0.305 | 68 |
| Neutrophil % | Trait anxiety | -0.111 | 0.773 | 0.980 | 68 | -0.706 | 0.09 | 0.240 | 68 |
| Neutrophil % | State anxiety | 0.212 | 0.586 | 0.980 | 68 | -0.935 | 0.018 | 0.188 | 68 |
| Eosinophil % | Depressive | -0.542 | 0.736 | 0.980 | 60 | 0.402 | 0.829 | 0.998 | 60 |
| Eosinophil % | Trait anxiety | 0.248 | 0.923 | 0.980 | 60 | -0.562 | 0.838 | 0.998 | 60 |
| Eosinophil % | State anxiety | -1.275 | 0.634 | 0.980 | 60 | -1.073 | 0.692 | 0.998 | 60 |
| Basophil % | Depressive | -0.864 | 0.905 | 0.980 | 60 | -0.409 | 0.963 | 0.998 | 60 |
| Basophil % | Trait anxiety | -1.759 | 0.879 | 0.980 | 60 | 3.174 | 0.808 | 0.998 | 60 |
| Basophil % | State anxiety | 0.605 | 0.960 | 0.980 | 60 | -4.122 | 0.748 | 0.998 | 60 |
| MLR | Depressive | -1.885 | 0.920 | 0.980 | 68 | -0.872 | 0.967 | 0.998 | 68 |
| MLR | Trait anxiety | 6.064 | 0.843 | 0.980 | 68 | 2.81 | 0.928 | 0.998 | 68 |
| MLR | State anxiety | 22.244 | 0.475 | 0.980 | 68 | -11.592 | 0.706 | 0.998 | 68 |
| NLR | Depressive | -2.54 | 0.225 | 0.980 | 68 | **-6.814** | **0.027** | 0.188 | 68 |
| NLR | Trait anxiety | -1.832 | 0.595 | 0.980 | 68 | -8.626 | 0.063 | 0.188 | 68 |
| NLR | State anxiety | 2.518 | 0.471 | 0.980 | 68 | **-9.375** | **0.036** | 0.188 | 68 |
| PLR | Depressive | 0.03 | 0.351 | 0.980 | 68 | 0.006 | 0.893 | 0.998 | 68 |
| PLR | Trait anxiety | 0.02 | 0.706 | 0.980 | 68 | 0 | 0.998 | 0.998 | 68 |
| PLR | State anxiety | 0.044 | 0.420 | 0.980 | 68 | -0.016 | 0.806 | 0.998 | 68 |
| Mitochondrial Respiratory Capacity (MRC) |  |  |  |  |  |  |  |  |  |
| Monocyte MRC | Depressive | 0.088 | 0.483 | 0.758 | 38 | -0.162 | 0.116 | 0.771 | 38 |
| Monocyte MRC | Trait anxiety | 0.097 | 0.638 | 0.841 | 38 | -0.081 | 0.604 | 0.798 | 38 |
| Monocyte MRC | State anxiety | 0.217 | 0.334 | 0.745 | 38 | -0.248 | 0.129 | 0.771 | 38 |
| Lymphocyte MRC | Depressive | **-0.258** | **0.023** | 0.386 | 58 | -0.150 | 0.258 | 0.798 | 58 |
| Lymphocyte MRC | Trait anxiety | -0.338 | 0.064 | 0.386 | 58 | -0.186 | 0.342 | 0.798 | 58 |
| Lymphocyte MRC | State anxiety | -0.234 | 0.205 | 0.616 | 58 | -0.189 | 0.318 | 0.798 | 58 |
| Neutrophil MRC | Depressive | -0.098 | 0.505 | 0.758 | 35 | -0.077 | 0.665 | 0.798 | 35 |
| Neutrophil MRC | Trait anxiety | -0.219 | 0.373 | 0.745 | 35 | -0.157 | 0.557 | 0.798 | 35 |
| Neutrophil MRC | State anxiety | -0.172 | 0.424 | 0.758 | 35 | -0.004 | 0.985 | 0.985 | 35 |
| PBMC AM MRC | Depressive | -0.079 | 0.669 | 0.841 | 65 | 0.123 | 0.533 | 0.798 | 65 |
| PBMC AM MRC | Trait anxiety | -0.05 | 0.868 | 0.868 | 65 | 0.149 | 0.603 | 0.798 | 65 |
| PBMC AM MRC | State anxiety | 0.056 | 0.855 | 0.868 | 65 | 0.073 | 0.794 | 0.841 | 65 |
| PBMC PM MRC | Depressive | -0.069 | 0.700 | 0.841 | 65 | 0.374 | 0.083 | 0.771 | 59 |
| PBMC PM MRC | Trait anxiety | -0.05 | 0.861 | 0.868 | 65 | 4.665 | 0.243 | 0.798 | 58 |
| PBMC PM MRC | State anxiety | 0.277 | 0.340 | 0.745 | 59 | 2.443 | 0.529 | 0.798 | 58 |
| Platelet MRC | Depressive | 0.162 | 0.087 | 0.389 | 31 | 0.058 | 0.552 | 0.798 | 31 |
| Platelet MRC | Trait anxiety | 0.282 | 0.057 | 0.386 | 31 | 0.043 | 0.741 | 0.834 | 31 |
| Platelet MRC | State anxiety | 0.209 | 0.142 | 0.510 | 31 | 0.058 | 0.629 | 0.798 | 31 |
| Respirometry |  |  |  |  |  |  |  |  |  |
| Monocytes Basal JATPox | Depressive | -0.019 | 0.646 | 0.845 | 65 | 0.037 | 0.440 | 0.949 | 65 |
| Monocytes Basal JATPox | Trait anxiety | 0.029 | 0.669 | 0.845 | 65 | 0.009 | 0.896 | 0.979 | 65 |
| Monocytes Basal JATPox | State anxiety | 0.097 | 0.145 | 0.497 | 65 | -0.016 | 0.820 | 0.979 | 65 |
| Monocytes Max JATPox | Depressive | 0.000 | 0.937 | 0.978 | 65 | 0.006 | 0.412 | 0.949 | 65 |
| Monocytes Max JATPox | Trait anxiety | 0.002 | 0.810 | 0.926 | 65 | 0.005 | 0.658 | 0.979 | 65 |
| Monocytes Max JATPox | State anxiety | 0.009 | 0.357 | 0.690 | 65 | 0.000 | 0.963 | 0.997 | 65 |
| Neutrophils Basal JATPox | Depressive | -0.001 | 0.998 | 0.998 | 61 | 0.115 | 0.600 | 0.960 | 61 |
| Neutrophils Basal JATPox | Trait anxiety | 0.415 | 0.214 | 0.627 | 61 | 0.079 | 0.806 | 0.979 | 61 |
| Neutrophils Basal JATPox | State anxiety | 0.203 | 0.540 | 0.810 | 61 | -0.220 | 0.474 | 0.949 | 61 |
| Neutrophils Max JATPox | Depressive | -0.047 | 0.395 | 0.690 | 61 | 0.040 | 0.571 | 0.960 | 61 |
| Neutrophils Max JATPox | Trait anxiety | -0.015 | 0.869 | 0.948 | 61 | 0.088 | 0.394 | 0.949 | 61 |
| Neutrophils Max JATPox | State anxiety | 0.028 | 0.759 | 0.911 | 61 | 0.013 | 0.898 | 0.979 | 61 |
| Lymphocytes Basal JATPox | Depressive | -0.197 | 0.045 | 0.217 | 64 | 0.071 | 0.533 | 0.960 | 64 |
| Lymphocytes Basal JATPox | Trait anxiety | -0.084 | 0.612 | 0.845 | 64 | 0.124 | 0.464 | 0.949 | 64 |
| Lymphocytes Basal JATPox | State anxiety | -0.136 | 0.403 | 0.690 | 64 | 0.135 | 0.402 | 0.949 | 64 |
| Lymphocytes Max JATPox | Depressive | **-0.052** | **0.011** | 0.176 | 60 | 0.000 | 0.997 | 0.997 | 60 |
| Lymphocytes Max JATPox | Trait anxiety | -0.026 | 0.446 | 0.713 | 60 | 0.010 | 0.754 | 0.979 | 60 |
| Lymphocytes Max JATPox | State anxiety | -0.039 | 0.244 | 0.627 | 60 | 0.009 | 0.761 | 0.979 | 60 |
| Platelets Basal JATPox | Depressive | 0.010 | 0.307 | 0.669 | 63 | 0.021 | 0.065 | 0.949 | 63 |
| Platelets Basal JATPox | Trait anxiety | **0.033** | **0.037** | 0.217 | 63 | 0.020 | 0.257 | 0.949 | 63 |
| Platelets Basal JATPox | State anxiety | 0.027 | 0.089 | 0.358 | 63 | 0.021 | 0.205 | 0.949 | 63 |
| Platelets Max JATPox | Depressive | 0.006 | 0.261 | 0.627 | 63 | 0.011 | 0.090 | 0.949 | 63 |
| Platelets Max JATPox | Trait anxiety | 0.022 | 0.015 | 0.176 | 63 | 0.010 | 0.323 | 0.949 | 63 |
| Platelets Max JATPox | State anxiety | 0.019 | 0.036 | 0.217 | 63 | 0.012 | 0.196 | 0.949 | 63 |

Moderation analyses were conducted in linear regression models between the variable and mood symptom of interest, with adversity group (high lifetime adversity or exposure to childhood adversity) as the covariate.

Next, we conducted these analyses based on levels of childhood adversity exposure measured using the childhood trauma questionnaire (CTQ, using cutoffs derived from ^63^). Of the entire MiSBIE cohort, 39% experienced no childhood trauma exposure, 61% experienced some childhood trauma exposure. Moderation analyses showed that childhood adversity exposure influenced the association between state anxiety symptoms and lymphocyte proportions (*b* = 1.10, *p* = 0.020, *p_adj_*= 0.19, Table 1) in healthy controls. That is, for healthy individuals exposed to childhood adversity, the positive association between state anxiety and lymphocyte proportions was stronger than in individuals who were not subject to childhood trauma exposure. In contrast, the opposite pattern was detected for the association between neutrophil proportions and state anxiety (*b* =-0.94, *p* = 0.018, *p_adj_*= 0.19), whereby the negative relation between these variables were significantly stronger in individuals who were subject to childhood trauma exposure than in individuals who were not. Further, childhood adversity levels also influenced the relation between NLR and depressive symptoms (*b* =-9.38, *p* = 0.027, *p_adj_* = 0.19, Table 1) and state anxiety symptoms (b =-6.81, p = 0.036, *p_adj_*= 0.19, Table 1). These results demonstrate that the above associations between mood symptoms and neutrophil measures are substantially more negative in the individuals who experienced childhood adversity than those who did not. Similar results including healthy controls and the MitoD group are presented in Table S2A).

Consistent with the moderation analysis results, among individuals who experienced childhood trauma, state anxiety symptoms were negatively associated with the proportion of neutrophils (*r* =-0.42, *p* = 0.005, *p_adj_* = 0.054, Fig 2E) and with NLR (*r* =-0.38, *p* = 0.010, *p_adj_* = 0.077), but positively associated with the proportion of lymphocytes (*r* = 0.34, *p* = 0.023, *p_adj_* =0.054). We also observed positive associations between the proportion of monocytes and depressive symptoms (*r* = 0.36, p = 0.018, *p_adj_* = 0.077), state anxiety (*r* = 0.44, p = 0.014, *p_adj_* = 0.054), and trait anxiety (*r* = 0.37, *p* = 0.003, *p_adj_* = 0.077) in participants who experienced childhood trauma (Fig 2D, F). In contrast, we did not detect significant associations among those who did not experience childhood trauma (Fig 2C). Similar patterns of association were again observed in the combined control + MitoD group, although the effect sizes were smaller, and fewer correlations reached significance (Fig S2C-D). Altogether, these findings suggest that the relation between immune cell proportion and depressive and anxiety symptoms may be influenced by early life adversity.

### Mitochondrial respiratory capacity and adversity

Next, we examined the associations between immune cell subtype and platelet mitochondrial biology and measures of adversity, anxiety, and depressive symptoms. Several measures of mitochondrial biology were combined into a single index of mitochondrial respiratory capacity (MRC, as described previously ^64^) in platelets, PBMCs (PBMCs AM collected in the morning, pre-psychosocial stress, fasted; PBMCs PM collected in the afternoon, post-psychosocial stress, fed), and isolated monocytes, lymphocytes, and neutrophils (Fig 3A).

**Figure 3.**
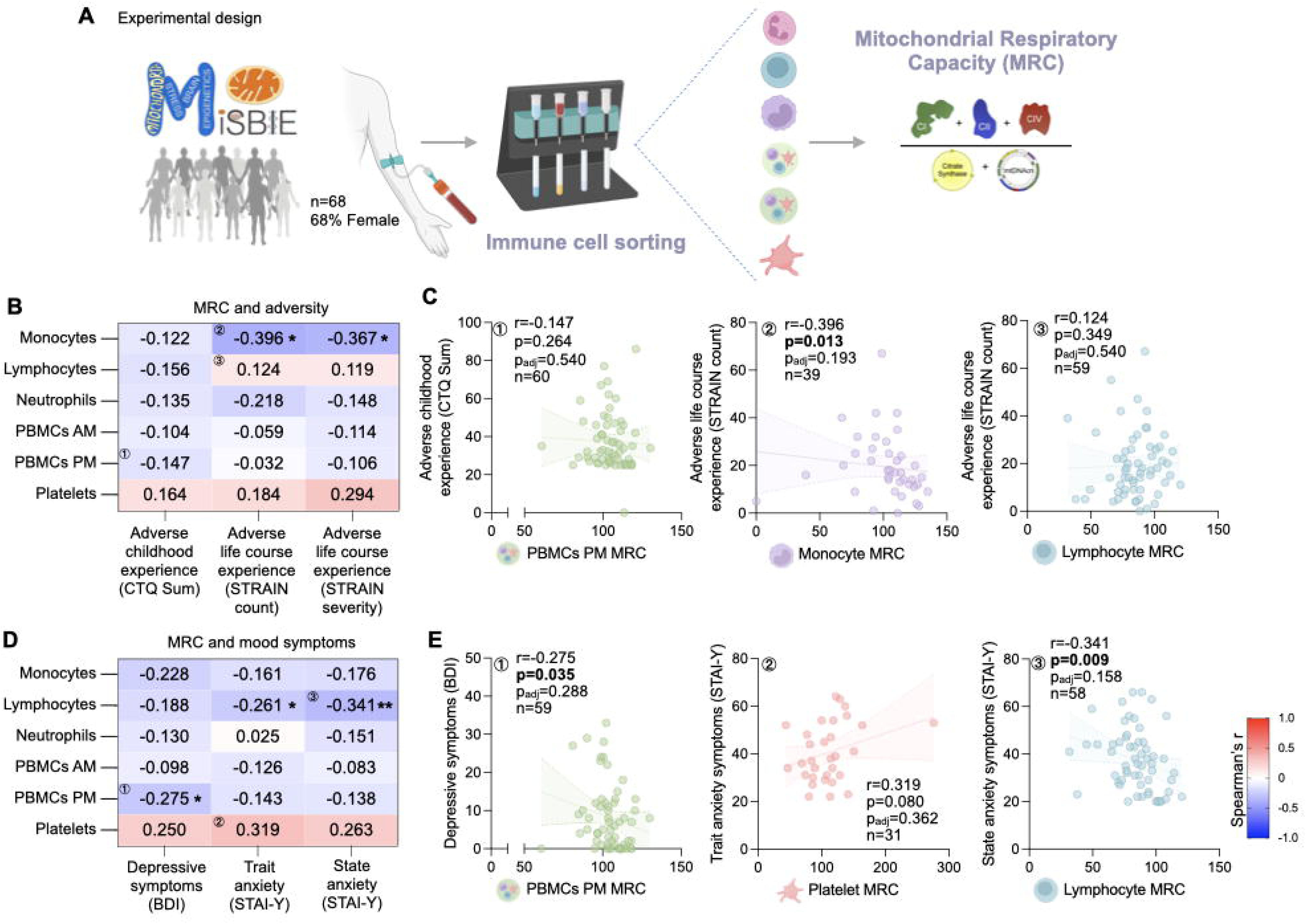
Associations between mitochondrial respiratory capacity (MRC) of different immune cell types with mood symptoms and psychosocial adversity. (A) Experimental design. Immune cells were isolated from blood collected from MiSBIE control participants to measure mitochondrial respiratory capacity (MRC). (B) Heatmap of Correlations between MRC of monocytes, lymphocytes, neutrophils, PBMCs (1/2), platelets and measures of adversity (CTQ, STRAIN Total Count, STRAIN Total Severity). n=32-66. (C) Scatterplots showing correlations between MRC and CTQ/STRAIN scores corresponding to the labeled comparisons on the heatmap. (D) Heatmap of correlations between MRC of monocytes, lymphocytes, neutrophils, PBMCs (1/2), platelets and measures of depression and anxiety (BDI, STAI-Trait, STAI-State). n=31-65. (E) Scatterplots showing correlations between MRC and BDI/STAI scores corresponding to the labeled comparisons on the heatmap. In scatterplots, linear regression lines shown for illustrative purposes only, all correlations are Spearman rank correlations. Effect size and p-values from Spearman rank correlation. *p<0.05, **p<0.01, ***p<0.001, ****p<0.0001. No correlations remained significant following Benjamini-Hochberg FDR correction.

In the context of mitochondrial bioenergetics, monocyte MRC was negatively correlated with lifetime stressor count (*r* =-0.40, *p* = 0.013, *p_adj_* = 0.19) and severity (*r* =-0.37, *p* = 0.021, *p_adj_* = 0.19) (Fig 3B-C). Overall, patterns of correlations suggested cell-type-specific relationships between MRC, individual MRC measures and lifetime adversity exposure (Fig 3B-C, Fig S3). Similar patterns of association were observed in the combined control + MitoD group (Fig S3). The results by lifetime adversity subscales are shown in (Fig S4).

### Mitochondrial respiratory capacity and mood symptoms

Regarding mood symptoms, we observed a pattern of negative correlations between MRC in immune cell subtypes and depression and both state and trait anxiety symptoms (Fig 3D). We found that MRC in lymphocytes was negatively correlated with state (*r* =-0.34, *p* = 0.009, *p_adj_* = 0.16) and trait (*r* =-0.26, *p* = 0.048, *p_adj_* = 0.29) anxiety symptoms (Fig 3D-E). In afternoon PBMCs, MRC also showed a negative association with depressive symptoms (*r* = - 0.28, *p* = 0.035, *p_adj_* = 0.29, Fig 3D-E). When examining individual mitochondrial biology indices, we observed negative associations between trait and state anxiety and Complex I (CI) activity in lymphocytes (Fig S5). Similar patterns of associations were observed in the combined control + MitoD group despite not consistently reaching significance (Fig S5).

### Mitochondrial respiratory capacity and mood symptoms by level of adversity

Given prior evidence that life adversity may modulate mitochondrial biology in immune cells ^34,65^, we investigated whether adversity also influences the association between MRC and mood symptoms by stratifying participants based on their level of lifetime stressor severity (below or above the participants’ median lifetime stressor severity score, as described above with the immune cell proportion analyses).

Moderation analyses revealed a significant effect of adversity group on the relation between lymphocyte MRC and depressive symptoms (*b* =-0.26, *p* = 0.023, *p_adj_* = 0.39, Table 1). Whereas those exposed to high lifetime adversity exhibited a negative correlation between lymphocyte MRC and depressive symptoms (*r* =-0.67, *p* = 0.0002, *p_adj_* = 0.0012, Fig 4B, E), those exposed to low lifetime adversity displayed weak, non-significant associations (Fig 4A). Lifetime adversity exposure also moderated the associations between mood symptoms and individual MRC measures in several cell subtypes (See Table S2B and Fig S6 for details). Notably, this was the case for the association between all six individual lymphocyte MRC variables and depressive symptoms (Table S2B). Individuals who experienced high lifetime adversity exhibited strong negative correlations between individual lymphocyte MRC variables and depressive symptoms (*r*’s =-0.51 to-0.67, *p*’s = 0.0002 to 0.027, *p_adj_’*s = 0.0012 to 0.088, Fig S6), whereas these associations were not found in those who experienced low lifetime adversity (Fig S6).

**Figure 4.**
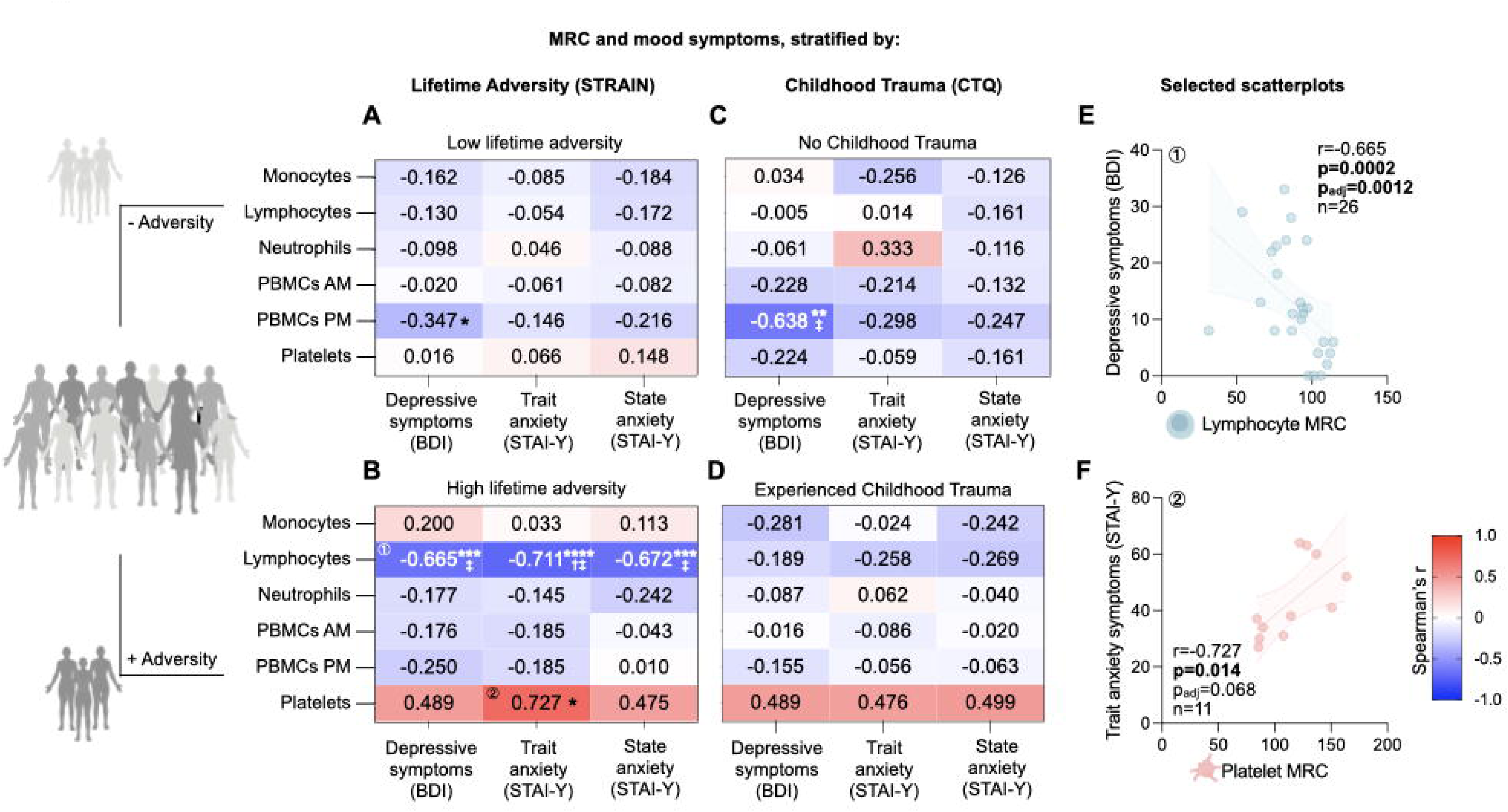
Associations between mitochondrial respiratory capacity (MRC) of different immune cell types with mood symptoms and psychosocial adversity, stratified by adversity exposure. (A) Heatmap of correlations between MRC of monocytes, lymphocytes, neutrophils, PBMCs (1/2), platelets and measures of depression and anxiety symptoms (BDI, STAI-Trait, STAI - State) in individuals who experienced low lifetime adversity (STRAIN severity score ≤44). n=20-38. (B) Heatmap of correlations between MRC of monocytes, lymphocytes, neutrophils, PBMCs (1/2), platelets and measures of depression and anxiety symptoms (BDI, STAI-Y Trait, STAI-Y-State) in individuals who have experienced high lifetime adversity (STRAIN severity score >44). n=11-27. (C) Heatmap of correlations between MRC of monocytes, lymphocytes, neutrophils, PBMCs (1/2), platelets and measures of depression and anxiety symptoms (BDI, STAI-Trait, STAI-State) in individuals who did not experience childhood trauma. n=13-20. (D) Heatmap of correlations between MRC of monocytes, lymphocytes, neutrophils, PBMCs (1/2), platelets and measures of depression and anxiety symptoms (BDI, STAI-Trait, STAI-State) in individuals who experienced childhood trauma. n=16-43. (E) Scatterplot of correlation between Lymphocyte MRC and depressive symptoms in controls in individuals who experienced high lifetime adversity. n=24. (F) Scatterplot of correlation between Platelet MRC and state anxiety symptoms in controls in individuals who experienced high lifetime adversity. n=10. In scatterplots, linear regression lines shown for illustrative purposes only, all correlations are Spearman rank correlations. Effect size and p-values from Spearman rank correlation. *p<0.05, **p<0.01, ***p<0.001, ****p<0.0001. Benjamini-Hochberg FDR-corrected ^†^p_adj_<0.05, ^‡^p_adj_<0.01, ^†‡^p_adj_<0.001, ^‡‡^p_adj_<0.0001.

In platelets, the effect of lifetime adversity group on the positive association between MRC and trait anxiety symptoms also approached significance (*b* = 0.28, *p* = 0.057, *p_adj_* = 0.39, Table 1), and in stratified analysis, platelet MRC exhibited a strong positive correlation with trait anxiety symptoms in individuals of the high lifetime adversity group (*r* = 0.73, p = 0.014, *p_adj_*= 0.068), but not in those of the low lifetime adversity group (Fig 4D, F).

Examining these associations based on childhood trauma exposure did not return significant interactions between childhood trauma and the relationship between lymphocyte MRC and all mood symptom measures (Table 2, Fig 4C-D). However, there was a significant moderating effect of childhood adversity on the relationship between lymphocyte CI and all mood symptoms (Fig S7, Table S2B). We also observed negative patterns of associations between lymphocyte MRC and mood symptoms in those who experienced childhood trauma although these did not reach significance. Further, for those with childhood trauma exposure, there was a negative association between lymphocyte CI activity and trait anxiety symptoms (*r* =-0.37, *p* = 0.022, *p_adj_* = 0.65), and state anxiety symptoms (*r* =-0.38, *p* = 0.020, *p_adj_* = 0.65) (Fig S7, Table S2B). We also observed several moderating effects of childhood trauma group on the relation between mood symptoms and other individual MRC measures of several cell subtypes (Fig S7, Table S2B). For example, at higher levels of life adversity, a significant negative association between depression and monocyte Complex II (CII) activity was found (*r* =-0.42, *p* = 0.047, *p_adj_* = 0.73, Fig S7, Table S2B). Similar results were observed with the combined control + MitoD group, although with weaker effect sizes (Fig S7, Table S2B). These results suggest reduced immune mitochondrial respiratory activity in white blood cells, particularly in lymphocytes, in the context of combined adversity exposure and mood symptoms.

### Respirometry, adversity, and mood symptoms

MRC is an indirect measure of the maximal possible respiration by the mitochondrial electron transport chain. To assess mitochondrial respiration in the context of living cells, we performed extracellular flux analysis of living immune cell subtypes and platelets from the same participants ^62^. Our primary analyses focused on ATP production rates derived from mitochondrial OxPhos under both basal (Basal *J*ATP_ox_) and maximal (Max *J*ATP_ox_) respiration. We also conducted additional exploratory analyses using additional parameters (Fig S8), including ATP production rates derived from cytosolic glycolysis under basal conditions (basal *J*ATP_gly_), total ATP cellular ATP production rate (basal *J*ATP_total_), the ratio of basal ATP production rate derived from mitochondrial OxPhos over basal ATP production rate derived from cytosolic glycolysis (Basal *J*ATP_ox/gly_), the percentage of basal oxygen consumption linked to ATP production (Coupling Efficiency), the ATP production rate derived from OxPhos under maximal respiratory activity without accounting for basal ATP production derived from OxPhos (Spare *J*ATP_ox_), and the Spare *J*ATP_ox_ expressed as a percentage of basal Basal *J*ATP_ox_ (Spare *J*ATP_ox_ Capacity). For a further explanation of these variables, see the methods, and for a schematic of these measures and what they represent, see Fig S8A.

No significant main effects were observed between either lifetime or childhood adversity measures or mood symptoms and immune cell subtype Basal *J*ATP_ox_ or Max *J*ATP_ox_ (Fig 5B-E). However, consistent with the MRC findings, in healthy participants, life adversity moderated the negative association between depressive symptoms and lymphocyte mitochondrial respiration parameters. Specifically, a significant interaction was observed for lymphocyte Max *J*ATP_ox_ (*b* =-0.052, *p* = 0.011, *p_adj_*= 0.18, Table 1), which was also found in the control + MitoD group (*b* = - 0.039, *p* = 0.018, *p_adj_*= 0.43, Table S2C). A significant interaction was observed for lymphocyte Basal *J*ATP_ox_ (*b* =-0.20, *p* = 0.045, *p_adj_* = 0.22, Table 1. Control + MitoD group results are shown in Table S2C). Stratified analyses revealed that no clear associations between mitochondrial respiration and depressive or anxiety symptoms were observed among individuals with low adversity exposure (Fig 6A). In contrast, among individuals with high adversity severity, we found a negative association between depressive symptoms and Basal *J*ATP_ox_ in lymphocytes (*r* =-0.45, *p* = 0.016, *p_adj_* = 0.19) and in monocytes (*r* =-0.45, *p* = 0.015, *p_adj_* = 0.19) (Fig 6B, E-F). Consistent with the MRC results, in the high lifetime adversity group, we found a trend for a negative association between lymphocyte Max *J*ATP_ox_ and depressive symptoms (*r* =-0.39, *p* = 0.052, *p_adj_* = 0.35, Fig 6B) that was not observed in the low adversity group.

**Figure 5.**
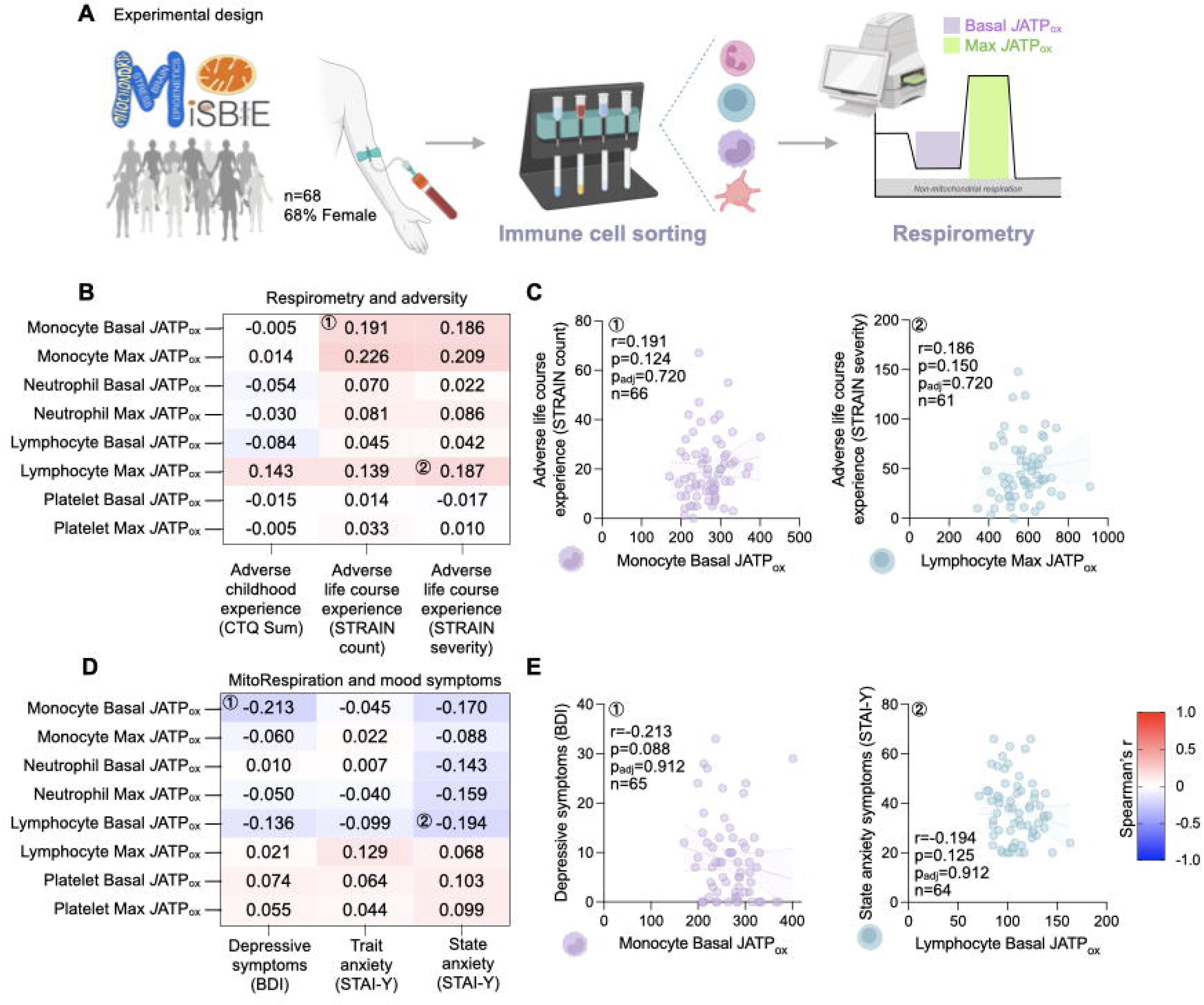
Associations between respirometry measures of different immune cell types with mood symptoms and psychosocial adversity. (A) Experimental design. Immune cells were isolated from blood collected from MiSBIE participants for respirometry. (B) Heatmap of Correlations between monocyte, lymphocyte and neutrophil respiration measures with measures of adversity (CTQ, STRAIN Total Count, STRAIN Total Severity). n=58-61. (C) Scatterplots showing correlations between monocyte, lymphocyte and neutrophil respiration measures with CTQ/STRAIN scores corresponding to the labeled comparisons on the heatmap. (D) Heatmap of correlations between monocyte, lymphocyte and neutrophil respiration measures with measures of depression and anxiety (BDI, STAI-Trait, STAI-State). n=57-60. (E) Scatterplots showing correlations between monocyte, lymphocyte and neutrophil respiration measures with BDI/STAI scores corresponding to the labeled comparisons on the heatmap. In scatterplots, linear regression lines shown for illustrative purposes only, all correlations are Spearman rank correlations. Effect size and p-values from Spearman rank correlation. *p<0.05, **p<0.01, ***p<0.001, ****p<0.0001. No correlations remained significant following Benjamini-Hochberg FDR correction.

**Figure 6.**
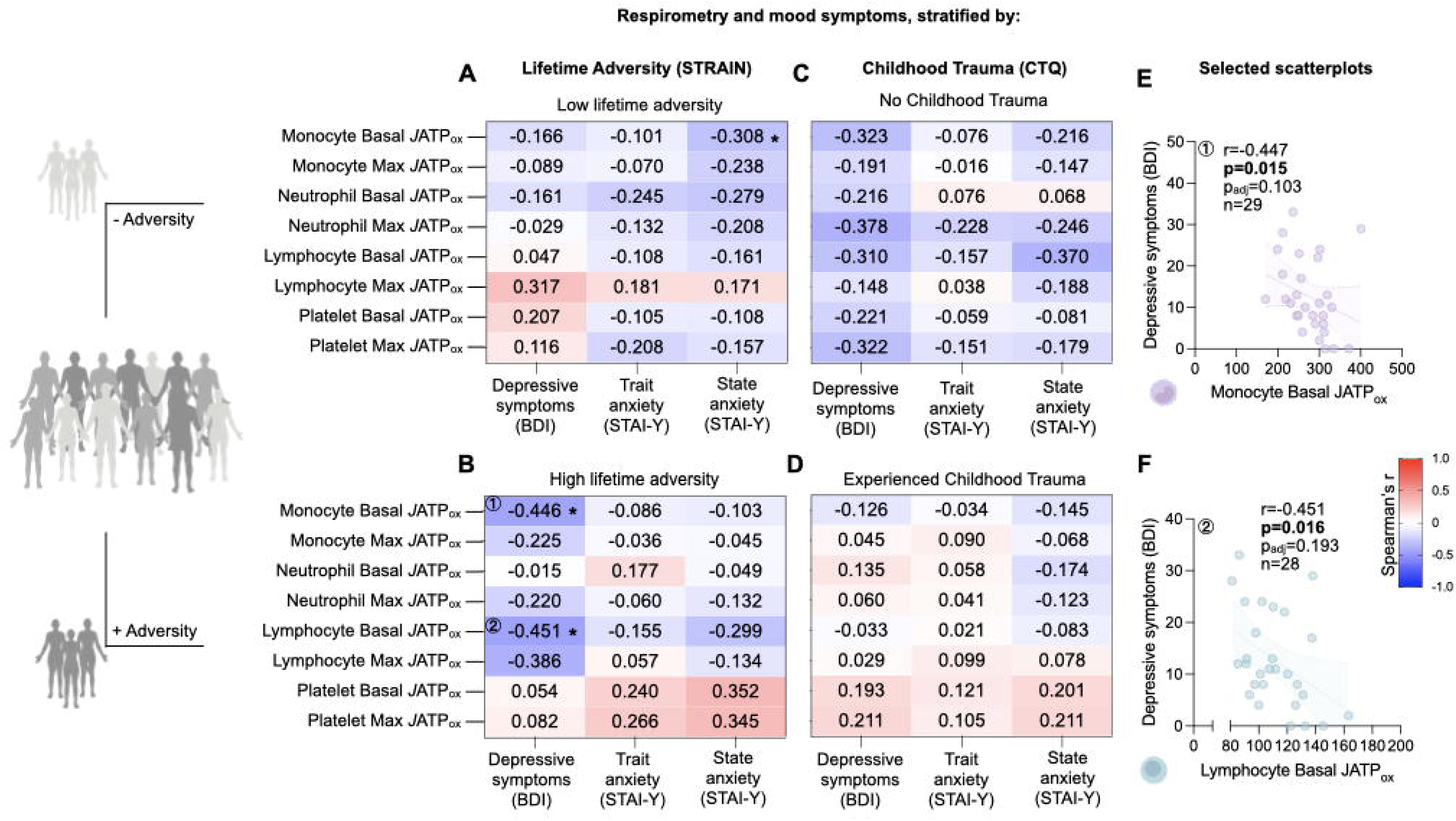
Associations between respirometry measures of different immune cell types with mood symptoms and psychosocial adversity, stratified by adversity exposure. (A) Heatmap of correlations between monocyte, lymphocyte and neutrophil respirometry measures with measures of depression and anxiety (BDI, STAI-Trait, STAI-State) in individuals who experienced low adversity (STRAIN severity score ≤50). n=33-35. (B) Heatmap of correlations between monocyte, lymphocyte and neutrophil respirometry measures with measures of depression and anxiety (BDI, STAI-Trait, STAI-State) in individuals who have experienced high adversity (STRAIN severity score >50). n=24-25. (C) Heatmap of correlations between monocyte, lymphocyte and neutrophil respirometry measures with measures of depression and anxiety (BDI, STAI-Trait, STAI-State) in controls who did not experience childhood trauma. n=18-20. (D) Heatmap of correlations between monocyte, lymphocyte and neutrophil respirometry measures with measures of depression and anxiety (BDI, STAI-Trait, STAI-State) in controls who experienced childhood trauma. n=39-40. Heatmap and select scatterplots of correlations between monocyte, lymphocyte and neutrophil respirometry measures with measures of depression and anxiety (BDI, STAI-Trait, STAI-State) in individuals who did not experience childhood trauma. n=30-32. (E) Scatterplot of correlation between Monocyte respirometry measures and depressive symptoms in controls in individuals who experienced high lifetime adversity. n=25. (F) Scatterplot of correlation between Lymphocyte respirometry measures and depressive symptoms in controls in individuals who experienced high lifetime adversity. n=25. In scatterplots, linear regression lines shown for illustrative purposes only, all correlations are Spearman rank correlations. Effect size and p-values from Spearman rank correlation. *p<0.05, **p<0.01, ***p<0.001, ****p<0.0001. No correlations remained significant following Benjamini-Hochberg FDR correction.

Consistent with the MRC results, moderation effects were observed for the association between anxiety symptoms and platelet respiration. Lifetime adversity significantly moderated the relation between trait anxiety symptoms and both platelet Basal *J*ATP_ox_ (*b* = 0.033, *p* = 0.037, *p_adj_* = 0.22, Table 1) and Max *J*ATP_ox_ (*b* = 0.022, p = 0.015, *p_adj_*= 0.18, Table 1). In individuals exposed to high lifetime adversity, we found trends of positive associations between platelet respiration and anxiety symptoms, that were not found in low lifetime adversity participants (Fig 6, S10). Similar associations with anxiety symptoms and the moderating effect of adversity exposure were observed for platelet Spare *J*ATP_ox_ and Basal *J*ATP_total_ (Table S2C, Fig S10).

Childhood adversity exposure was not a significant moderator of the relationship between mood symptoms with Basal *J*ATP_ox_ and Max *J*ATP_ox_ across cell types (Table 2, Fig 6C, D, S11).

Altogether, these Seahorse-based mitochondrial bioenergetics findings broadly converge with the biochemical MRC results. These results, collected via two complementary approaches, suggest that lymphocyte mitochondrial respiratory capacity is negatively associated with depressive symptoms, whereas platelet mitochondrial respiratory capacity trends positively with anxiety symptoms in individuals exposed to higher levels of adversity.

## 3. Discussion

In this study, we investigated for the first time how lifetime adversity and mood symptoms were related to mitochondrial biology across specific immune cell subtypes. Our findings, summarized in Figure 7, reveal that adversity exposure and mood symptoms are associated with cell-type-specific changes in immune cell proportions and mitochondrial biology. Stronger associations were observed for individuals who experienced greater lifetime adversity. These findings suggest that lifetime adversity may lead to immune and mitochondrial adaptations, that could ultimately contribute or interact with anxiety and depressive symptoms.

**Figure 7.**
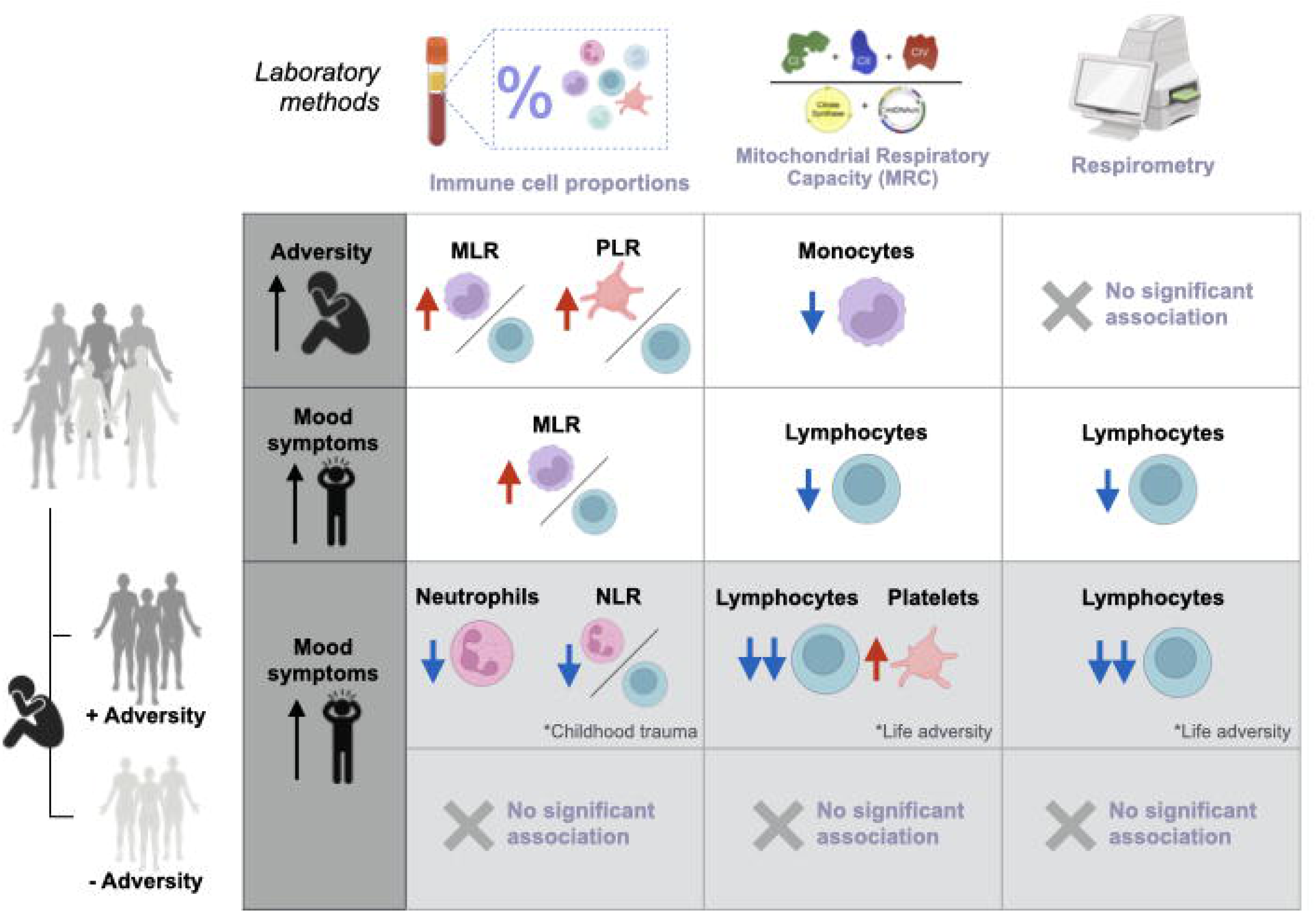
Summary of results from this study. Adversity was positively correlated with MLR, PLR and negatively correlated with Monocyte MRC. Mood symptoms were positively correlated with MLR and negatively correlated with lymphocyte MRC and respirometry. Mood symptoms were negatively correlated with neutrophil proportion, NLR, lymphocyte MRC and respirometry in individuals exposed to adversity.

Consistent with prior research ^12,13,66^, our results show alterations in immune cell and platelet proportions in relation to adversity and mood symptoms. Specifically, greater adversity was associated with higher MLR and PLR, and mood symptoms were related to higher MLR.

These findings are consistent with prior studies reporting elevated monocytes and reduced lymphocytes proportions with depression ^26,27^. Importantly, childhood adversity amplified negative associations between neutrophil proportion and mood symptoms. Further, there was a significant positive relation between mood symptoms and monocyte proportion in individuals exposed to childhood trauma. This is consistent with previous reports connecting adversity to immune changes, with immune gene expression changes found in monocytes ^18,67–71^. Taken together, these findings underscore the relevance of understanding the dynamic interplay between adversity, mood, and immune function across development and into adulthood.

Regarding mitochondrial bioenergetics, we found that higher adversity was associated with lower monocyte in-vitro enzymatic MRC but not live cell respirometry measures, warranting further investigation. We also observed consistent negative associations between both MRC measures and respiration in lymphocytes and mood disorder symptoms, and these associations were stronger in individuals who experienced greater levels of lifetime adversity. This suggests that the reduction in lymphocyte mitochondrial bioenergetics in response to adversity may be relevant in adversity’s role of increasing the risk of experiencing mood symptoms. High mitochondrial respiration in lymphocytes is associated with quiescent or naïve functional phenotypes in the absence of immune activation ^72^. Therefore, lower lymphocyte mitochondrial respiration may reflect immune cell activation and elevated inflammation ^73–75^, which is prevalent in both adversity and depression severity ^76^.

In platelets, we observed positive associations between anxiety symptoms and both MRC and respirometry results, particularly in individuals that experienced high lifetime adversity. This aligns with previous research demonstrating elevated CI mRNA levels and activity, as well as CI/CS ratio and CIV activity in bipolar disorder ^51,77^. This is also consistent with work showing greater platelet CI activity is associated with higher schizophrenia and depressive symptoms ^49^ severity. However, our results contrast with prior research demonstrating decreased platelet mitochondrial respiration in depression ^78^. Although data on the role of platelet respiration in anxiety is limited, platelet activation has been shown to be affected by stress, and in turn platelet activation has been shown to impact mitochondrial respiration ^79^. Our findings provide rationale to further investigate the association between adversity exposure, platelet bioenergetics, and mood symptoms.

We did not find any consistent association between adversity measures or mood symptoms and PBMC mitochondrial bioenergetics. Prior studies have shown reductions of PBMC mitochondrial bioenergetics to be linked with greater psychosocial stress and adversity exposure ^36,80,81^, particularly with regard to threat and deprivation dimensions of early life adversity ^82^, and reduction of mitochondrial respiration in PBMCs ^50^ of depressed patients. In contrast, other reports have shown increased PBMC respiration with adversity exposure ^34^. These mixed results, alongside the cell-type-specific nature of associations observed in this study, underscores the importance of studying mitochondrial function in isolated immune cell subtypes beyond mixed populations. The divergence of associations between mood symptoms and mitochondrial bioenergetics between lymphocytes and platelets may reflect their distinct roles in immune and systemic responses to stress. Lymphocytes, as adaptive immune cells, may be more sensitive to chronic stress-induced mitochondrial adaptations and negatively regulate mitochondrial respiration under stressful conditions. In contrast, platelets, which play key roles in hemostasis and inflammation and are upregulated during stress, may increase mitochondrial activity as a compensatory mechanism under stress conditions.

Although our study provides important new insights into the underlying biology of lifetime adversity exposure and mood symptoms, several limitations should be acknowledged. First, the cross-sectional and retrospective design of this research precludes causal inferences about the directionality of these associations. Thus, longitudinal studies are needed to determine whether mitochondrial adaptations in specific immune cells precede or result from mood disorders. Second, although we assessed symptoms of depression and anxiety, these associations have yet to be examined in individuals clinically diagnosed with mood disorders across immune cell types and mitochondrial variables.

Additionally, specific lymphocyte cell subtypes (e.g. B vs. T cells) are known to possess further distinct mitochondrial respiratory levels. This can influence overall lymphocyte mitochondrial respiration and functions ^58,83,84^. Mood disorders have also been shown to relate to differential changes in circulating lymphocyte cell types ^85–87^, therefore circulating proportions of lymphocyte subtypes may also result in differential relationships between lymphocyte metabolism and mood disorders. As a result, reduced respiratory capacity observed in the present study may reflect not only mitochondrial alterations within cells but also shifts in circulating lymphocyte subtype composition. Future studies should account for lymphocyte subset composition.

A further limitation of this study is the large number of associations performed given the number of immune and mitochondrial variables available in our dataset. While we focused on unadjusted p-values in our discussion and interpretation of these results, an important limitation to this study is that many of these results did not remain significant following FDR correction. That being said, we see patterns of results that are convergent across different analyses, with both MRC and Respirometry measures demonstrating similar results in the analyses between bioenergetics, mood and adversity metrics, particularly in lymphocytes. Further, within individual MRC and Respirometry measures, we also see consistency in the directionality of associations. This convergence could suggest that the p-values, while few in significance, may reflect a true signal rather than scattered false positives.

Despite these limitations, our study provides novel insights into cell-type-specific associations between mitochondrial biology, adversity, and mood disorders. By highlighting the potential role of lifetime stressor and adversity exposure in shaping immune mitochondrial biology, this study contributes to a growing body of literature emphasizing the importance of bioenergetics in mental health ^40,88^. Alongside a number of studies that have established the relationship between chronic stressor exposure and recalibrations in mitochondrial biology across tissues ^33,89,90^, this study further supports the hypothesis that stressor exposure may become biologically embedded through changes in mitochondrial biology ^62^. We further build on this prior research by examining the impact of adversity experienced throughout lifetime and childhood. The moderating effects of life adversity on mitochondrial biology suggest that targeted stress-reduction intervention may be effective in mitigating mitochondrial dysregulation and its potential downstream effects on mood—an effect that has been found for other biological outcomes ^91^. The distinct patterns of associations observed between immune cells underscore the need for further studies on immune cell types to elucidate the molecular mechanisms linking adversity, immune mitochondrial biology, and mood disorders.

In conclusion, our study demonstrates that life adversity and mood symptoms are associated with distinct adaptations in immune cell proportions and mitochondrial biology. The stronger associations observed for individuals exposed to greater lifetime adversity provides important new insight into how life stress may get under the skin to potentially affect health. This finding, along with others ^12,40,85–88^, point to an interplay between bioenergetics and immune cell function that could potentially manifest in the development of mood disorders.

## 4. Methods

### Participant Recruitment

Participants aged 18 to 60 were recruited to the Mitochondrial Stress, Brain Imaging, and Epigenetics (MiSBIE) study in adherence to the directives outlined by the New York State Psychiatric Institute IRB protocol #7424, ClinicalTrials.gov NCT04831424. Participant recruitment details and detailed procedures for the 2-day onsite study visit can be found in previously published reports ^62^. Recruitment occurred at various research sites, including Columbia University Irving Medical Center, across clinical partners in the United States and Canada, through national consortia (e.g., NAMDC, UMDF), and Columbia’s RecruitMe site. A total of 105 participants were included in the present study, with a subset of 68 healthy controls and 37 individuals with genetically defined mitochondrial diseases (MitoD). All enrolled participants provided written informed consent, authorizing their participation in the investigative procedures and the dissemination of findings.

### Participants

Eligibility criteria for healthy controls included: willingness to provide saliva samples and have a venous catheter installed for blood collection during their hospital visit, as well as the absence of pregnancy (in females). Exclusion criteria for healthy controls included cognitive deficits to provide informed consent, occurrences of seasonal infections in the four weeks prior to the study, Raynaud’s syndrome, ongoing involvement in therapeutic or exercise-related trials on ClinicalTrials.gov, the presence of metal inside or outside the body, and claustrophobia impeding participation in magnetic resonance imaging (MRI) procedures.

Eligibility criteria for MitoD patients included a genetic diagnosis of either (i) the m.3243A > G point mutation, with or without mitochondrial encephalopathy, lactic acidosis, and stroke-like episodes (MELAS), or (ii) a single, large-scale mtDNA deletion—often associated chronic progressive external ophthalmoplegia (CPEO) or Kearns-Sayre syndrome (KSS). Exclusion criteria were similar to those of healthy controls, with additional items excluding individuals using ongoing steroid therapy (oral dexamethasone, prednisone, or similar), steroid use, and current diagnosis of neoplastic disease.

### Procedures

Blood was drawn in a fasted state at ∼10 am on Day 1 to obtain immune cell subtypes, PBMCs 1, and platelets, to measure cell proportions (One 3mL K_2_EDTA tube), and mitochondrial bioenergetics (Five 8.5mL ACD-A tubes (BD-364606). Next, participants completed a series of psychological tasks, behavioral questionnaires, and medical examinations, which were part of additional MiSBIE procedures not examined here. In the afternoon, participants were asked to complete a socio-evaluative speech task. A final blood draw was performed two hours after the speech task for the assessment of MRC in PBMCs (PBMCs 2). At the end of day 1, participants completed questionnaires assessing life adversity exposure. On day 2, participants also completed a series of surveys to capture mood symptoms and childhood adversity measures.

### Depressive symptoms, anxiety, and adversity

Depressive symptoms were assessed using the commonly used Beck Depression Inventory II (BDI) ^92^. Trait and state anxiety symptoms were assessed using the State and Trait Anxiety Inventory (STAI-Y) ^93^. Lifetime stressor count and severity were assessed using the well-validated Stress and Adversity Inventory for Adults (STRAIN; see http://www.strainsetup.com). The complete STRAIN was administered, but only the six core lifetime stressor exposure scores and time-limited count and severity subscales were analyzed. The “high lifetime adversity” group was defined as those who had a STRAIN lifetime stressor severity score above the median of the entire MiSBIE cohort. The “low lifetime adversity” group was comprised of participants with a STRAIN lifetime stressor severity score below the median of the entire MiSBIE cohort. Childhood adversity measures were assessed using the Childhood Trauma Questionnaire (CTQ) ^94^, which included five subscales: physical, emotional, sexual abuse, as well as emotional and physical neglect. These subscales were summed to produce a CTQ total score. Established cutoffs were used to assess childhood trauma in any specific subscale ^63^. If the participant experienced trauma in any subscale, they were categorized in the “experienced childhood trauma” group. Otherwise, they were categorized in the “no childhood trauma” group. Mean CTQ total scores were 46 in the “experienced childhood trauma” group and 25 in the “no childhood trauma” group.

### Complete blood count (CBC) with differential

To measure proportions of lymphocytes, monocytes, neutrophils, basophils, eosinophils and platelets, a complete blood count (CBC) with differential was performed from blood collected in the morning fasting state at 10 am. This was conducted with a Sysmex XN in an FDA-approved, and CLIA-certified laboratory. Additional information about the assay can be found at https://www.testmenu.com/nyphcolumbia/Tests/624139.

### Cell type isolation for Mitochondrial Respiratory Capacity Measurements and Respirometry

Mitochondrial enzyme activity and mtDNAcn measurements were quantified in monocytes, lymphocytes, neutrophils, and PBMCs. These were isolated using a double Ficoll gradient (platelets and PBMCs) and magnetic bead labelled antibodies (monocytes, lymphocytes and neutrophils) immediately after sample collection. Detailed blood processing methods can be found at ^62,64^. Briefly, samples were collected via a central venous catheter in ACD-A tubes (BD-364606) at 10 am in the morning for monocyte, lymphocyte, neutrophil, PBMC AM, and platelet collection, and at 4 pm post-stress task for PBMC PM collection. Samples were then centrifuged at 500g for 5min at room temperature with the brakes off.

Platelet isolation was performed using 3ml of morning plasma processed and suspended with prostaglandin I2 (PGI2) (Cayman Chemicals, #18220). Platelets were quantified with a turbidity assay for the first 28 participants, estimated using the formula described in ^95^, and the remaining samples were quantified using a Bicinchoninic acid (BCA) assay (Bioworld #20831001) for whole protein content. Samples for Mitochondrial Respiratory Capacity measurements were stored in liquid nitrogen until sample homogenization and enzyme activity assays were run.

PBMC isolation was performed using morning and afternoon blood. Whole blood was added to 5mL HBSS (Gibco, #14175103) in a 15ml tube. The diluted blood was slowly layered over 4mL of preloaded Histopaque 1077 (Sigma, #10771) in a 15ml conical tube. The tube was then spun at 400g for 30min at room temperature with brakes off. The cell layer was transferred to a 15ml conical tube with HBSS, final volume 15ml. The tube was centrifuged at 500g for 10 minutes to pellet cells, the supernatant was discarded, and cells were resuspended with HBSS to 15ml for washing. The tube was then centrifuged twice at 200g for 10 minutes at room temperature, discarding the supernatant each time and resuspending the pellet in 15 mL HBSS. After the third wash, the supernatant was discarded, and cells were resuspended in 1ml of HBSS. Next, 10μl of suspension was mixed with 10μl of trypan blue and counted using the Countess II automated cell counter (Invitrogen). Total cell count in the suspension was estimated via the cell count and recorded. Samples were aliquoted before spinning at 2000g for 2min, having the supernatant aspirated, and stored in a-80C freezer before transfer into liquid nitrogen.

Immune cell subtypes (monocytes, lymphocytes and neutrophils) were isolated from 3ml of buffy coat after platelet isolation. Whole blood was overlaid on a double gradient of 10ml Histopaque 1077 and 10ml Histopaque 1119 in two 50mL tubes, then tubes were spun at 700g for 30min at room temperature, with the brakes off. The middle mononuclear (MCN) cell layer was collected for monocyte and lymphocyte isolation. The polymorphonuclear (PMN) cell layer was collected for neutrophil isolation. Cell layers were pooled in respective 50ml tubes, each with 5ml HBSS, followed by centrifugation and resuspension, in 1ml of HBSS/BSA (0.5% BSA in HBSS) (Sigma, #A3733). Cells were aliquoted, centrifuged, pelleted, and resuspended before the addition of 250ul HBSS/BSA. Monocyte, neutrophil and lymphocytes were isolated using magnetic bead labelled antibodies and MACs separator columns (Miltenyi Biotech, #130042401). Monocytes were isolated from the MCN tube using magnetic bead labelled CD14 antibody (Miltenyi Biotec, #130050201), and the neutrophils were isolated from the PMN tube using magnetic bead labelled CD15 antibody (Miltenyi Biotec, #130046601). Lymphocytes were isolated from the flowthrough after the monocyte isolation. The flowthrough was centrifuged at 700g for 40 seconds. The resulting cell pellet was resuspended in HBSS/BSA using magnetic beads labeled CD61 (Miltenyi Biotec, #130051101) and CD235 (Miltenyi Biotec, #130050501) antibodies.

Cell counts were performed using an automated cell counter (Countess II, Invitrogen). Samples for Mitochondrial Respiratory Capacity measurements were stored in liquid nitrogen until enzymatic assays were run. Meanwhile, samples for Seahorse *in vivo extracellular flux* measurements were resuspended in XF Media containing no pH buffers and supplemented with 5.5 mM glucose (Gibco, #15023021), 1 mM sodium pyruvate (Gibco, #11360070), 1 mM L-glutamine (Gibco, #25030081), 50 ug/ml uridine (Sigma, #U6381-5G), and 10 mM palmitate (Sigma, #P9767-5G) conjugated to 1.7 mM BSA without fatty acids (Sigma, #A3733-50G).

### Mitochondrial Respiratory Capacity Measurements: Sample Homogenization

Isolated immune cell samples were homogenized as previously described ^57^ in 500μl of homogenization buffer (1mM EDTA, 50mM Triethanolamine) with two tungsten beads (Qiagen Cat#69997) and pre-chilled racks in a Tissue Lyser (Qiagen, Cat# 85300). Cells were lysed with two rounds of 30 cycles/s for 1min, with incubation of samples at 4°C in ice for 5min between rounds. Mouse reference samples for batch correction across plates were prepared by homogenizing mouse tissue with 180μl homogenization buffer per gram tissue, using the same procedure as the cells, but with an additional three freeze/thaw cycles.

### Mitochondrial enzyme measurements

Mitochondrial enzyme activity measurements were completed as previously described ^57,64^ with minor modifications. Briefly, Complex I (CI) activity was determined by measuring the change in absorbance at 600 nm over 10min, in a 100uM potassium phosphate reaction buffer (pH 7.5) containing 2mM EDTA, 3.mg/ml BSA, 0.25mM potassium cyanide, 10μM decylubuquinone, 100μM DCIP, 200μM NADH, and 0.4μM antimycin A. Non-specific activity was detected in the presence of 500uM rotenone and 200uM piericidin A. Complex II (CII) activity was measured as the change in absorbance at 600nm over 15min, in 100mM potassium phosphate reaction buffer (pH 7.5) with mM EDTA, 1mg/ml BSA, 4μM rotenone, 1mM succinate, 0.25mM potassium cyanide, 100μM decylubuquinone, 100μM DCIP, 200μM ATP, and 0.4μM antimycin A. Non-specific activity was measured in the presence of malonate (5 mM). Complex IV (CIV) activity was measured as the change in absorbance at 550nm over 20min for lymphocytes, neutrophils, monocytes, and platelets, and 10min for PBMCs, in 100mM potassium phosphate reaction buffer (pH 7.5) containing 0.1% n-dodecylmaltoside and 50μM of purified reduced cytochrome c. Spontaneous cytochrome c oxidation was determined by measuring change in absorbance without cell homogenate. Citrate Synthase (CS) activity was determined by measuring the change in absorbance at 412nm over 8min, in reaction buffer (200mM Tris, pH 7.4) containing 0.2mM acetyl-CoA 0, 0.2mM 5,5’-dithiobis-(2-nitrobenzoic acid) (DTNB), 0.55mM oxaloacetic acid, and 0.1% Triton X-100. Non-specific activity was measured as the change in absorbance without oxaloacetate in the reaction buffer.

Enzymatic assays were performed in 96-well plates and recorded on a Spectramax M2 (Spectramax Pro 6, Molecular Devices), using the following volumes of homogenate: Complex I: 15μl, COX and SDH: 20μl, and CS: 10μl. Total and non-specific activities for each enzyme were assessed in triplicate at 30°C. For CIV, the number of replicates of spontaneous cytochrome c oxidation measure was equal to the number of samples assessed. Activities were determined using the change in absorbance over time, where the trace was linear and maximal. OD was transformed to enzymatic activity, using the following molar extinction coefficients: DTNB – 13.6Lmol^-1^cm^-1^, reduced cytochrome c – 29.5Lmol^-1^cm^-1^, DCIP – 16.3Lmol^-1^cm^-1^.

Specific activities were calculated by taking the average of total activities and subtracting the average non-specific/spontaneous activities. Any specific activity that was calculated to be <0 was deemed undetectable and replaced by 0. Samples that had a CV greater than the cut-off CVs (CS, CI, SDH – 15%, COX – 35% for PBMCs and 30% for all other cell types) within the replicates and that were one standard deviation from the mean of the replicates were excluded. If any datapoints were excluded due to non-linear traces, the average of the remaining measures was taken.

Batch correction for each enzyme assay was performed by dividing enzyme activities per plate by a correction factor calculated using mouse activity reference samples on each plate, normalized to the average of the reference samples across all plates. This was averaged across reference samples on each given plate to produce a correction factor. Following batch correction, enzymatic activities of immune cell types were normalized to nDNA content quantified via qPCR (method below); while platelets were normalized to protein concentration quantified via a BCA assay kit (Bioworld #20831001). Biologically implausible outliers were then identified as samples that were more than 3 times the interquartile range (IQR) higher than the 3^rd^ quartile, and 3 times the IQR lower than the 1^st^ quartile. Detailed normalization and data cleaning methods are described at ^96^.

### mtDNAcn quantification

mtDNA copy number (mtDNAcn) and nuclear DNA (nDNA) were measured via qPCR as previously described ^57^, with minor modifications, and are described in detail elsewhere (see ^64^). 20μl enzymatic activity homogenate was lysed in 180μl lysis buffer (6% Tween20 (Sigma #P1379), 114 mM Tris-HCl pH 8.5 (Sigma #T3253), and 200 µg/mL Proteinase K (Thermofisher #AM2548) for 16h at 55°C, followed by 95°C for 10min, and maintained at 4°C until used for qPCR if qPCR was done within 24h of lysis, or otherwise stored at-80°C until qPCR. qPCR reactions were performed in two plates of triplicates in 384-well qPCR plates with 8μl of lysate and 12μl of master mix, cycled in a QuantStudio 7 flex qPCR instrument (Applied Biosystems, Cat# 4485701) with the following conditions: 50°C for 2min, 95°C for 20s, followed by 40 cycles of 95°C for 1s, and 60°C for 20min (total run time: 40min).

Taqman chemistry was used to simultaneously quantify mitochondrial and nuclear amplicons for two distinct primer pairs, ND1/B2M and COX1/RNaseP. Master Mixes for each primer pair included TaqMan Universal Master mix fast (Life Technologies #4444964), 300nM of primers and 100nM probe. ND1-Fwd: GAGCGATGGTGAGAGCTAAGGT, ND1-Rev: CCCTAAAACCCGCCACATCT, Probe: HEX-CCATCACCCTCTACATCACCGCCC-3IABkFQ. B2M-Fwd: CCAGCAGAGAATGGAA AGTCAA, B2M-Rev: TCTCTCTCCATTCTTCAGTAAGTCAACT, Probe: FAM-ATGTGTCTGGGT TTCATCCATCCGACA-3IABkFQ). COX1-Fwd: CTAG CAGGTGTCTCCTCTATCT, COX1-Rev: GAGAAGTAGGACTGCTGTGATTAG, Probe: HEX-TGCC ATAACCCAATACCAAACGCC-3IABkFQ. RNaseP-Fwd: AGATTTGGACCTGCGAGCG, RNaseP-Rev: GAGCGGCTGTCTCCACAAGT, Probe: FAM-TTCTGACCTGAAGGCTCTGCGCG-3IABkFQ.

### mtDNAcn calculation

mtDNAcn was calculated as 2 2, where ΔCt was the average mtDNA Ct subtracted from the average nDNA Ct. The final mtDNAcn was calculated by averaging the mtDNAcn calculated from each primer pair, unless sample values deviated more than 20% from the mean deviation between the primer pair for the plate. Samples with such deviation were systematically checked for obvious amplification failures. If an amplification failure was found, the mtDNAcn value calculated with the failed primer was replaced with a predicted value based on a plate-specific linear regression model.

### mtDNA in platelets

Platelets do not have nDNA, so here we report mtDNA content normalized to protein concentration derived from the linearized mtDNA Ct divided by protein concentration of each sample. Protein concentrations in the enzymatic activity homogenates were measured via BCA assay kit (Bioworld #20831001).

### In vivo extracellular flux measurements

Oxygen consumption rate (OCR) and extracellular acidification rate (ECAR) were measured using a Seahorse XFe96 Analyzer (Agilent Technologies) as described in ^62^. In brief, isolated immune cells were resuspended in XF media containing no pH buffers and supplemented with 5.5 mM glucose (Gibco, #15023021), 1 mM sodium pyruvate (Gibco, #11360070), 1 mM L-glutamine (Gibco, #25030081), 50 ug/ml uridine (Sigma, #U6381-5G), and 10 mM palmitate (Sigma, #P9767-5G) conjugated to 1.7 mM BSA without fatty acids (Sigma, #A3733-50G). Cells were seeded at a density of 250,000 cells per well in six wells of a poly-D-lysine-coated Seahorse cell culture plate. Plates were centrifuged (pulsed spin) to promote cell attachment and incubated at 37 °C without CO₂ for 1 hour. The Seahorse instrument was programmed to assess OCR and ECAR after the sequential addition of oligomycin (Sigma, #75351, final concentration: 1 mM), FCCP (Sigma, #C2920, final concentration: 2 mM), Rotenone (Sigma, #R8875, final concentration: 1 mM) and Antimycin A (Sigma #A8674, final concentration: 1 mM). After each run, OCR and ECAR measurements were normalized with either a protein concentration (Mi001-014) or using image-based cell counts (Mi016-110). OCR and ATP production rates derived from mitochondrial oxidative phosphorylation and glycolysis were calculated as previously described ^97,98^.

We focused on the following variables in our main analyses: 1) Basal *J*ATP_ox_ (pmol ATP/min) - ATP production rate derived from mitochondrial OxPhos under basal conditions; 2) Max *J*ATP_ox_ (pmol ATP/min) - ATP production rate derived from OxPhos under maximal respiratory activity. Additionally, we conducted supplementary analyses for the following variables: 1) Basal *J*ATP_gly_ (pmol ATP/min) - ATP production rate derived from cytosolic glycolysis under basal conditions; 2) Basal *J*ATP_tot_ (pmol ATP/min) - ATP production rate derived from both glycolysis and OxPhos under basal conditions (algebraic sum), can be interpreted as basal ATP consumption rate as this is measured under steady state; 3) Basal *J*ATP_ox/gly_ – Ratio between Basal ATP production rate derived from mitochondrial OxPhos over basal ATP production rate derived from cytosolic glycolysis, which informs us on which pathway the cell relies the most to generate the ATP consumed. Additional bioenergetic parameters relative to mitochondrial flexibility include: 4) Coupling Efficiency (%) - Percentage of basal oxygen consumption rate that is linked to ATP production; 5) Spare *J*ATP_ox_ (pmol ATP/min) - ATP production rate derived from OxPhos under maximal respiratory activity without accounting for the basal *J*ATP_ox_; 6) Spare *J*ATP_ox_ capacity (%) - Spare *J*ATP_ox_ expressed as a percentage of basal *J*ATP_ox_.

### Statistical Analyses

All statistical analyses were done using GraphPad Prism (Version 10.2.0) and R (version 4.3.2). Differences between healthy participants and MitoD were assessed in GraphPad Prism using Mann-Whitney tests for continuous variables and Fisher’s exact tests for categorical variables. Spearman’s *r* (*r*) analyses were used to test the associations between cell-proportion indices, cell type-specific mitochondrial features, and depression/anxiety and adversity measures in GraphPad Prism. Moderation analyses were conducted in R with adversity group or childhood trauma exposure as moderators to assess their influence on the associations between mood symptoms and cell-proportion indices or cell type-specific mitochondrial features. Stratified analyses by level of adversity are also presented. Given the exploratory nature of these analyses, we focused on uncorrected p-values (*p*), however Benjamini-Hochberg FDR-corrected p-values (*p_adj_*) are also reported in the results. FDR corrections were performed per figure using R. Some analyses performed in the main table and figures are also included in the supplemental tables and figures for reference. Main analyses presented in the main figures were restricted to the healthy control group. Analyses applied to the full MiSBIE cohort (healthy control + MitoD) are presented in Figs S1, S3-11 and Table S2A-C. Analyses restricted to the MitoD group, although underpowered, are reported in Tables S3 and S4.

## Data Availability

Data available upon reasonable request.

## Acknowledgements

We are grateful to the entire MiSBIE study team (www.picardlab.org/MiSBIE), and the participants and their families who made this study possible.

## Funding

The work of the authors was supported by NIH grants R01MH122706 and the Baszucki Group to M.P., Wharton Fund to C.T. and M.P. G.M.S. was supported by grant #OPR21101 from the California Governor’s Office of Planning and Research/California Initiative to Advance Precision Medicine. The findings and conclusions in this article are those of the authors and do not necessarily represent the views or opinions of these organizations, which had no role in designing or planning this study; in collecting, analyzing, or interpreting the data; in writing the article; or in deciding to submit this article for publication.

## Conflict of interests

The authors declare no conflicts of interest with respect to this work.

## Notes

### Competing Interest Statement

The authors have declared no competing interest.

### Author Declarations

Participants aged 18 to 60 were recruited to the Mitochondrial Stress, Brain Imaging, and Epigenetics (MiSBIE) study in adherence to the ethical directives outlined by the IRB of New York State Psychiatric Institute, protocol #7424, ClinicalTrials.gov NCT04831424.

### Summary of Updates

A correction was made to Figure 7; the illustration has been changed to reflect that a finding related to platelets was made using the method 'MRC', not 'respirometry'.

## References

1 Bandelow, B. & Michaelis, S. Epidemiology of anxiety disorders in the 21st century. Dialogues Clin Neurosci 17, 327–335 (2015). 10.31887/DCNS.2015.17.3/bbandelow

2 Kessler, R. C. et al. Lifetime prevalence and age-of-onset distributions of mental disorders in the World Health Organization’s World Mental Health Survey Initiative. World Psychiatry 6, 168–176 (2007).

3 Hertzman, C. The biological embedding of early experience and its effects on health in adulthood. Ann N Y Acad Sci 896, 85–95 (1999). 10.1111/j.1749-6632.1999.tb08107.x

4 Richter-Levin, G. & Xu, L. How could stress lead to major depressive disorder? IBRO reports 4, 38–43 (2018).

5 Young, E. A., Abelson, J. L., Curtis, G. C. & Nesse, R. M. Childhood adversity and vulnerability to mood and anxiety disorders. Depression and anxiety 5, 66–72 (1997).

6 Hassel, S., McKinnon, M. C., Cusi, A. M. & MacQueen, G. M. An overview of psychological and neurobiological mechanisms by which early negative experiences increase risk of mood disorders. Journal of the Canadian Academy of Child and Adolescent Psychiatry 20, 277 (2011).

7 Liu, R. T. Childhood adversities and depression in adulthood: Current findings and future directions. Clinical psychology: science and practice 24, 140 (2017).

8 Wang, Z.-y., et al. Dimensional early life adversity and anxiety symptoms: A network analysis and longitudinal study. Child Abuse & Neglect 160, 107201 (2025).

9 Juruena, M. F., Eror, F., Cleare, A. J. & Young, A. H. The Role of Early Life Stress in HPA Axis and Anxiety. Adv Exp Med Biol 1191, 141–153 (2020). 10.1007/978-981-32-9705-0_9

10 Ulrich-Lai, Y. M. & Herman, J. P. Neural regulation of endocrine and autonomic stress responses. Nature reviews neuroscience 10, 397–409 (2009).

11 Marcolongo-Pereira, C. et al. Neurobiological mechanisms of mood disorders: Stress vulnerability and resilience. Frontiers in behavioral neuroscience 16, 1006836 (2022).

12 Elwenspoek, M. M., Kuehn, A., Muller, C. P. & Turner, J. D. The effects of early life adversity on the immune system. Psychoneuroendocrinology 82, 140–154 (2017).

13 Etzel, L. et al. Immune cell dynamics in response to an acute laboratory stressor: a within-person between-group analysis of the biological impact of early life adversity. Stress 25, 347–356 (2022).

14 Apsley, A. T. et al. Investigating the effects of maltreatment and acute stress on the concordance of blood and DNA methylation methods of estimating immune cell proportions. Clinical epigenetics 15, 33 (2023).

15 Kuhlman, K. R., Horn, S. R., Chiang, J. J. & Bower, J. E. Early life adversity exposure and circulating markers of inflammation in children and adolescents: A systematic review and meta-analysis. Brain, behavior, and immunity 86, 30–42 (2020).

16 Segerstrom, S. C. & Miller, G. E. Psychological stress and the human immune system: a meta-analytic study of 30 years of inquiry. Psychological bulletin 130, 601 (2004).

17 Chiang, J. J., Lam, P. H., Chen, E. & Miller, G. E. Psychological stress during childhood and adolescence and its association with inflammation across the lifespan: A critical review and meta-analysis. Psychological Bulletin 148, 27 (2022).

18 Bower, J. E. et al. Childhood maltreatment and monocyte gene expression among women with breast cancer. Brain, behavior, and immunity 88, 396–402 (2020).

19 Cole, S. W. et al. Transcriptional modulation of the developing immune system by early life social adversity. Proc Natl Acad Sci U S A 109, 20578–20583 (2012). 10.1073/pnas.1218253109

20 Marie-Mitchell, A. & Cole, S. W. Adverse childhood experiences and transcriptional response in school-age children. Dev Psychopathol 34, 875–881 (2022). 10.1017/s095457942000187x

21 Schwaiger, M. et al. Altered Stress-Induced Regulation of Genes in Monocytes in Adults with a History of Childhood Adversity. Neuropsychopharmacology 41, 2530–2540 (2016). 10.1038/npp.2016.57

22 Stein, M., Miller, A. H. & Trestman, R. L. Depression, the immune system, and health and illness: findings in search of meaning. Archives of General Psychiatry 48, 171–177 (1991).

23 Maes, M. et al. Leukocytosis, monocytosis and neutrophilia: hallmarks of severe depression. Journal of psychiatric research 26, 125–134 (1992).

24 Irwin, M. et al. Reduction of immune function in life stress and depression. Biological psychiatry 27, 22–30 (1990).

25 Casiani, R. I. Á. et al. Monocyte profiles and their association with depression severity and functional disability. Journal of Psychiatric Research 184, 272–278 (2025).

26 Daray, F. M. et al. Decoding the inflammatory signature of the major depressive episode: insights from peripheral immunophenotyping in active and remitted condition, a case-control study. Transl Psychiatry 14, 254 (2024). 10.1038/s41398-024-02902-2

27 Sørensen, N. V. et al. Immune cell composition in unipolar depression: a comprehensive systematic review and meta-analysis. Molecular Psychiatry 28, 391–401 (2023).

28 Wen, X. et al. Sex-specific association of peripheral blood cell indices and inflammatory markers with depressive symptoms in early adolescence. Journal of Affective Disorders 362, 134–144 (2024).

29 Surtees, P. et al. Association of depression with peripheral leukocyte counts in EPIC-Norfolk—role of sex and cigarette smoking. Journal of psychosomatic research 54, 303– 306 (2003).

30 Rezin, G. T. et al. Inhibition of mitochondrial respiratory chain in brain of rats subjected to an experimental model of depression. Neurochem Int 53, 395–400 (2008). 10.1016/j.neuint.2008.09.012

31 Weger, M. et al. Mitochondrial gene signature in the prefrontal cortex for differential susceptibility to chronic stress. Scientific reports 10, 18308 (2020).

32 Hunter, R. G. et al. Stress and corticosteroids regulate rat hippocampal mitochondrial DNA gene expression via the glucocorticoid receptor. Proceedings of the National Academy of Sciences 113, 9099–9104 (2016).

33 Trumpff, C. et al. Psychosocial experiences are associated with human brain mitochondrial biology. Proceedings of the National Academy of Sciences 121, e2317673121 (2024).

34 Boeck, C. et al. Inflammation in adult women with a history of child maltreatment: The involvement of mitochondrial alterations and oxidative stress. Mitochondrion 30, 197– 207 (2016). 10.1016/j.mito.2016.08.006

35 Boeck, C. et al. Targeting the association between telomere length and immuno-cellular bioenergetics in female patients with Major Depressive Disorder. Sci Rep 8, 9419 (2018). 10.1038/s41598-018-26867-7

36 Gumpp, A. M. et al. Childhood maltreatment is associated with changes in mitochondrial bioenergetics in maternal, but not in neonatal immune cells. Proc Natl Acad Sci U S A 117, 24778–24784 (2020). 10.1073/pnas.2005885117

37 Kurkinen, K. et al. The associations between metabolic profiles and sexual and physical abuse in depressed adolescent psychiatric outpatients: an exploratory pilot study. Eur J Psychotraumatol 14, 2191396 (2023). 10.1080/20008066.2023.2191396

38 Andreazza, A. C., Shao, L., Wang, J. F. & Young, L. T. Mitochondrial complex I activity and oxidative damage to mitochondrial proteins in the prefrontal cortex of patients with bipolar disorder. Arch Gen Psychiatry 67, 360–368 (2010). 10.1001/archgenpsychiatry.2010.22

39 Ben-Shachar, D. & Karry, R. Neuroanatomical pattern of mitochondrial complex I pathology varies between schizophrenia, bipolar disorder and major depression. PLoS One 3, e3676 (2008). 10.1371/journal.pone.0003676

40 Holper, L., Ben-Shachar, D. & Mann, J. J. Multivariate meta-analyses of mitochondrial complex I and IV in major depressive disorder, bipolar disorder, schizophrenia, Alzheimer disease, and Parkinson disease. Neuropsychopharmacology 44, 837–849 (2019). 10.1038/s41386-018-0090-0

41 Konradi, C. et al. Molecular evidence for mitochondrial dysfunction in bipolar disorder. Arch Gen Psychiatry 61, 300–308 (2004). 10.1001/archpsyc.61.3.300

42 Sun, X., Wang, J. F., Tseng, M. & Young, L. T. Downregulation in components of the mitochondrial electron transport chain in the postmortem frontal cortex of subjects with bipolar disorder. J Psychiatry Neurosci 31, 189–196 (2006).

43 Whatley, S. A., Curti, D. & Marchbanks, R. M. Mitochondrial involvement in schizophrenia and other functional psychoses. Neurochem Res 21, 995–1004 (1996). 10.1007/bf02532409

44 Gardner, A. & Boles, R. G. Mitochondrial energy depletion in depression with somatization. Psychother Psychosom 77, 127–129 (2008). 10.1159/000112891

45 Gardner, A. et al. Alterations of mitochondrial function and correlations with personality traits in selected major depressive disorder patients. J Affect Disord 76, 55–68 (2003). 10.1016/s0165-0327(02)00067-8

46 Kuffner, K. et al. Major Depressive Disorder is Associated with Impaired Mitochondrial Function in Skin Fibroblasts. Cells 9 (2020). 10.3390/cells9040884

47 de Sousa, R. T. et al. Lithium increases leukocyte mitochondrial complex I activity in bipolar disorder during depressive episodes. Psychopharmacology (Berl) 232, 245–250 (2015). 10.1007/s00213-014-3655-6

48 Fernström, J. et al. Blood-based mitochondrial respiratory chain function in major depression. Transl Psychiatry 11, 593 (2021). 10.1038/s41398-021-01723-x

49 Fišar, Z. et al. Activities of mitochondrial respiratory chain complexes in platelets of patients with Alzheimer’s disease and depressive disorder. Mitochondrion 48, 67–77 (2019). 10.1016/j.mito.2019.07.013

50 Karabatsiakis, A. et al. Mitochondrial respiration in peripheral blood mononuclear cells correlates with depressive subsymptoms and severity of major depression. Transl Psychiatry 4, e397 (2014). 10.1038/tp.2014.44

51 Akarsu, S. et al. Mitochondrial complex I and III mRNA levels in bipolar disorder. J Affect Disord 184, 160–163 (2015). 10.1016/j.jad.2015.05.060

52 Hofstra, B. M., Hoeksema, E. E., Kas, M. J. & Verbeek, D. S. Cross-species analysis uncovers the mitochondrial stress response in the hippocampus as a shared mechanism in mouse early life stress and human depression. Neurobiol Stress 31, 100643 (2024). 10.1016/j.ynstr.2024.100643

53 Tyrka, A. R. et al. Alterations of Mitochondrial DNA Copy Number and Telomere Length With Early Adversity and Psychopathology. Biol Psychiatry 79, 78–86 (2016). 10.1016/j.biopsych.2014.12.025

54 Chacko, B. K. et al. Methods for defining distinct bioenergetic profiles in platelets, lymphocytes, monocytes, and neutrophils, and the oxidative burst from human blood. Laboratory Investigation 93, 690–700 (2013). 10.1038/labinvest.2013.53

55 Kramer, P. A., Ravi, S., Chacko, B., Johnson, M. S. & Darley-Usmar, V. M. A review of the mitochondrial and glycolytic metabolism in human platelets and leukocytes: Implications for their use as bioenergetic biomarkers. Redox Biology 2, 206–210 (2014). 10.1016/j.redox.2013.12.026

56 Maianski, N. A. et al. Functional characterization of mitochondria in neutrophils: a role restricted to apoptosis. Cell Death & Differentiation 11, 143–153 (2004). 10.1038/sj.cdd.4401320

57 Rausser, S. et al. Mitochondrial phenotypes in purified human immune cell subtypes and cell mixtures. Elife 10, e70899 (2021).

58 Weiss, S. L. et al. Influence of Immune Cell Subtypes on Mitochondrial Measurements in Peripheral Blood Mononuclear Cells From Children with Sepsis. Shock 57 (2022).

59 Dhabhar, F. S. Effects of stress on immune function: the good, the bad, and the beautiful. Immunologic research 58, 193–210 (2014).

60 Herbert, T. B. & Cohen, S. Stress and immunity in humans: a meta-analytic review. Psychosomatic medicine 55, 364–379 (1993).

61 Schedlowski, M. et al. Changes of natural killer cells during acute psychological stress. Journal of clinical immunology 13, 119–126 (1993).

62 Kelly, C. et al. A Platform to Map the Mind-Mitochondria Connection and the Hallmarks of Psychobiology: The MiSBIE Study. Trends in Endocrinology & Metabolism 35, 884–901 (2024).

63 Nakajima, M. et al. Validation of childhood trauma questionnaire-short form in Japanese clinical and nonclinical adults. Psychiatry Research Communications 2, 100065 (2022). 10.1016/j.psycom.2022.100065

64 Liu, C. C. et al. Immune cell mitochondrial phenotypes are largely preserved in mitochondrial diseases and do not reflect disease severity. bioRxiv, 2025.2001. 2018.633635 (2025).

65 Boeck, C. et al. The association between cortisol, oxytocin, and immune cell mitochondrial oxygen consumption in postpartum women with childhood maltreatment. Psychoneuroendocrinology 96, 69–77 (2018). 10.1016/j.psyneuen.2018.05.040

66 Elwenspoek, M. M. et al. Proinflammatory T cell status associated with early life adversity. The Journal of Immunology 199, 4046–4055 (2017).

67 Kuhlman, K. R., Cole, S. W., Tan, E. N., Swanson, J. A. & Rao, U. Childhood maltreatment and immune cell gene regulation during adolescence: Transcriptomics highlight non-classical monocytes. Biomolecules 14, 220 (2024).

68 Kuhlman, K. R., Tan, E. N., Cole, S. W. & Rao, U. Differential immune profiles in the context of chronic stress among childhood adversity-exposed adolescents. *Brain*, Behavior, and Immunity 127, 183–192 (2025).

69 Miller, G. E., Cohen, S. & Ritchey, A. K. Chronic psychological stress and the regulation of pro-inflammatory cytokines: a glucocorticoid-resistance model. Health Psychol 21, 531–541 (2002). 10.1037//0278-6133.21.6.531

70 Slavich, G. M. & Cole, S. W. The Emerging Field of Human Social Genomics. Clin Psychol Sci 1, 331–348 (2013). 10.1177/2167702613478594

71 Slavich, G. M., Mengelkoch, S. & Cole, S. W. Human social genomics: Concepts, mechanisms, and implications for health. Lifestyle Med (Hoboken) 4 (2023). 10.1002/lim2.75

72 Ganeshan, K. & Chawla, A. Metabolic regulation of immune responses. Annu Rev Immunol 32, 609–634 (2014). 10.1146/annurev-immunol-032713-120236

73 Frauwirth, K. A. et al. The CD28 signaling pathway regulates glucose metabolism. Immunity 16, 769–777 (2002).

74 Ganeshan, K. & Chawla, A. Metabolic regulation of immune responses. Annual review of immunology 32, 609–634 (2014).

75 Wang, R. et al. The transcription factor Myc controls metabolic reprogramming upon T lymphocyte activation. Immunity 35, 871–882 (2011).

76 Bekhbat, M., Ulukaya, G. B., Bhasin, M. K., Felger, J. C. & Miller, A. H. Cellular and immunometabolic mechanisms of inflammation in depression: Preliminary findings from single cell RNA sequencing and a tribute to Bruce McEwen. Neurobiol Stress 19, 100462 (2022). 10.1016/j.ynstr.2022.100462

77 Zvěřová, M. et al. Disturbances of mitochondrial parameters to distinguish patients with depressive episode of bipolar disorder and major depressive disorder. Neuropsychiatric disease and treatment, 233–240 (2019).

78 Hroudová, J., Fišar, Z., Kitzlerová, E., Zvěřová, M. & Raboch, J. Mitochondrial respiration in blood platelets of depressive patients. Mitochondrion 13, 795–800 (2013). 10.1016/j.mito.2013.05.005

79 Grichine, A. et al. The fate of mitochondria during platelet activation. Blood Advances 7, 6290–6302 (2023).

80 Picard, M. et al. A mitochondrial health index sensitive to mood and caregiving stress. Biological psychiatry 84, 9–17 (2018).

81 Wu-Chung, E. L. et al. Mitochondrial Health, Physical Functioning, and Daily Affect: Bioenergetic Mechanisms of Dementia Caregiver Well-Being. Psychosom Med 86, 512– 522 (2024). 10.1097/psy.0000000000001312

82 Cleveland, S. et al. Early Life Adversity and Mitochondrial Function: Comparing Cumulative Risk and Dimensional Models of Adversity. Biological Psychiatry 10.1016/j.biopsych.2026.04.006

83 Liang, H. et al. Elucidating the mitochondrial function of murine lymphocyte subsets and the heterogeneity of the mitophagy pathway inherited from hematopoietic stem cells. Frontiers in immunology 13, 1061448 (2022).

84 Liu, Z. et al. Mitochondria-related parameters of lymphocyte subsets can distinguish different disease stages in patients with HBV infection. Scientific Reports 15, 21008 (2025).

85 Li, S. et al. Circulating T-cell subsets discrepancy between bipolar disorder and major depressive disorder during mood episodes: A naturalistic, retrospective study of 1015 cases. CNS Neurosci Ther 30, e14361 (2024). 10.1111/cns.14361

86 Rizzo, L. B. et al. An immunological age index in bipolar disorder: A confirmatory factor analysis of putative immunosenescence markers and associations with clinical characteristics. International Journal of Methods in Psychiatric Research 27, e1614 (2018).

87 Grosse, L. et al. Circulating cytotoxic T cells and natural killer cells as potential predictors for antidepressant response in melancholic depression. Restoration of T regulatory cell populations after antidepressant therapy. Psychopharmacology 233, 1679–1688 (2016).

88 Daniels, T. E., Olsen, E. M. & Tyrka, A. R. Stress and Psychiatric Disorders: The Role of Mitochondria. Annu Rev Clin Psychol 16, 165–186 (2020). 10.1146/annurev-clinpsy-082719-104030

89 Huang, Q. et al. The energetic stress cytokine GDF15 is elevated in the context of chronic and acute psychosocial stress. bioRxiv, 2024.2004.2019.590241 (2025). 10.1101/2024.04.19.590241

90 Duchowny, K. A. et al. Childhood adverse life events and skeletal muscle mitochondrial function. Science Advances 10, eadj6411 (2024).

91 Shields, G. S., Spahr, C. M. & Slavich, G. M. Psychosocial Interventions and Immune System Function: A Systematic Review and Meta-analysis of Randomized Clinical Trials. JAMA Psychiatry 77, 1031–1043 (2020). 10.1001/jamapsychiatry.2020.0431

92 Beck, A. T., Ward, C. H., Mendelson, M., Mock, J. & Erbaugh, J. An inventory for measuring depression. Arch Gen Psychiatry 4, 561–571 (1961). 10.1001/archpsyc.1961.01710120031004

93 Spielberger, C., Gorsuch, R., Lushene, R., Vagg, P. R. & Jacobs, G. Manual for the State-Trait Anxiety Inventory (Form Y1 – Y2). Vol. IV (1983).

94 Bernstein, D. P., Ahluvalia, T., Pogge, D. & Handelsman, L. Validity of the Childhood Trauma Questionnaire in an adolescent psychiatric population. J Am Acad Child Adolesc Psychiatry 36, 340–348 (1997). 10.1097/00004583-199703000-00012

95 Walkowiak, B., Kesy, A. & Michalec, L. Microplate reader--a convenient tool in studies of blood coagulation. Thromb Res 87, 95–103 (1997). 10.1016/s0049-3848(97)00108-4

96 Liu, C. C. et al. Immune cell mitochondrial phenotypes are largely preserved in mitochondrial diseases and do not reflect disease severity. Neurology: Genetics 12, e200343 (2026).

97 Desousa, B. R. et al. Calculation of ATP production rates using the Seahorse XF Analyzer. EMBO Rep 24, e56380 (2023). 10.15252/embr.202256380

98 Mookerjee, S. A., Gerencser, A. A., Nicholls, D. G. & Brand, M. D. Quantifying intracellular rates of glycolytic and oxidative ATP production and consumption using extracellular flux measurements. J Biol Chem 292, 7189–7207 (2017). 10.1074/jbc.M116.774471

