## Supplemental Figures S1-S11 for "Lifetime adversity exposure, mood symptoms, and immune mitochondrial bioenergetics"

#### Figure S1

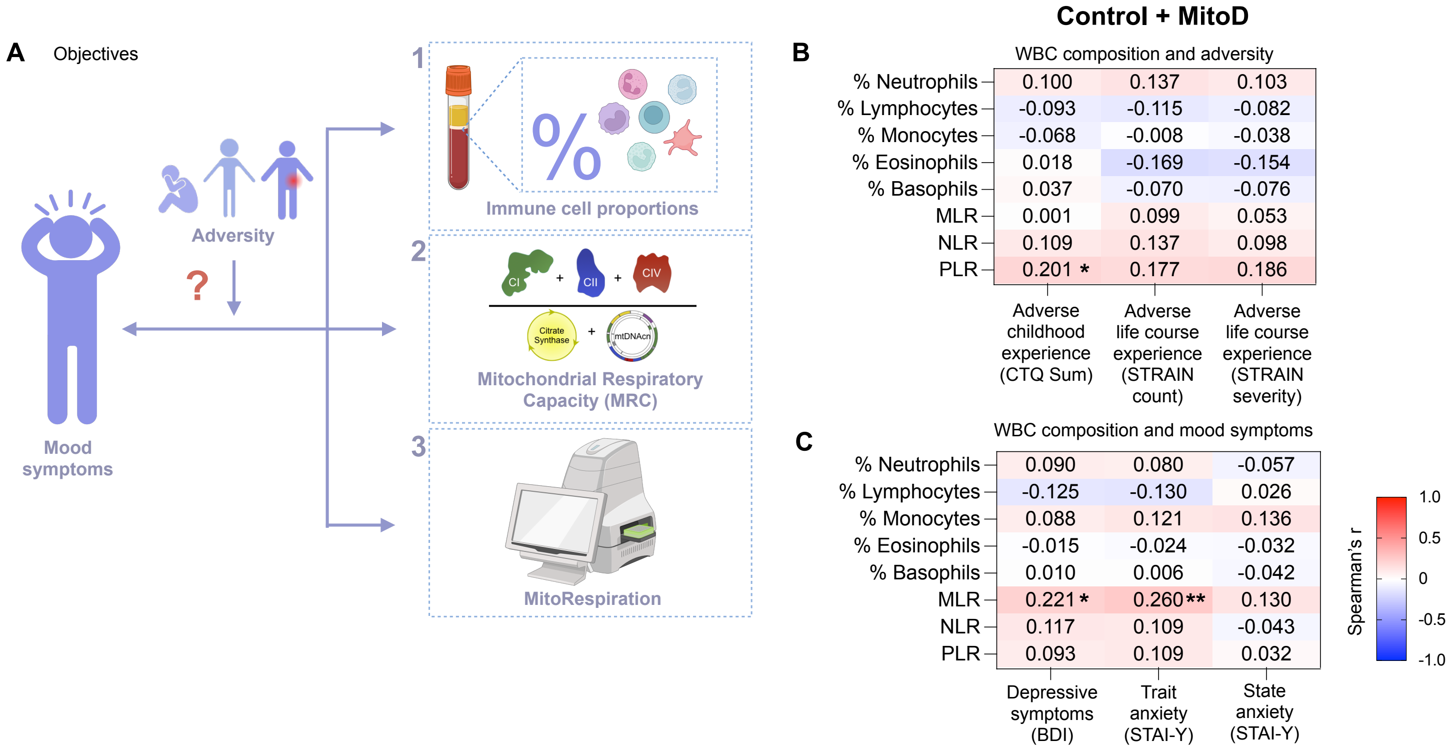


**Figure S1. Correlations between white blood cell composition with psychosocial measures in controls only**

(A) Objectives. We hypothesize that the relationship between immune cell proportions, MRC and MitoRespiration with psychiatric symptoms may be affected by adversity exposure.

(B) Heatmap of Correlations between WBC percentages and ratios with measures of adversity (CTQ, STRAIN Total Count, STRAIN Total Severity). n=105.

(C) Heatmap of correlations between white blood cell population (WBC) percentages and monocyte lymphocyte ratio (MLR), neutrophil lymphocyte ratio (NLR), platelet lymphocyte ratio (PLR) with measures of depression and anxiety (BDI, STAI-Trait, STAI-State). n=104-106.

Effect size and p-values from Spearman rank correlation. *p<0.05, **p<0.01, ***p<0.001, ****p<0.0001. No correlations remained significant following Benjamini-Hochberg FDR correction.

### Figure S2

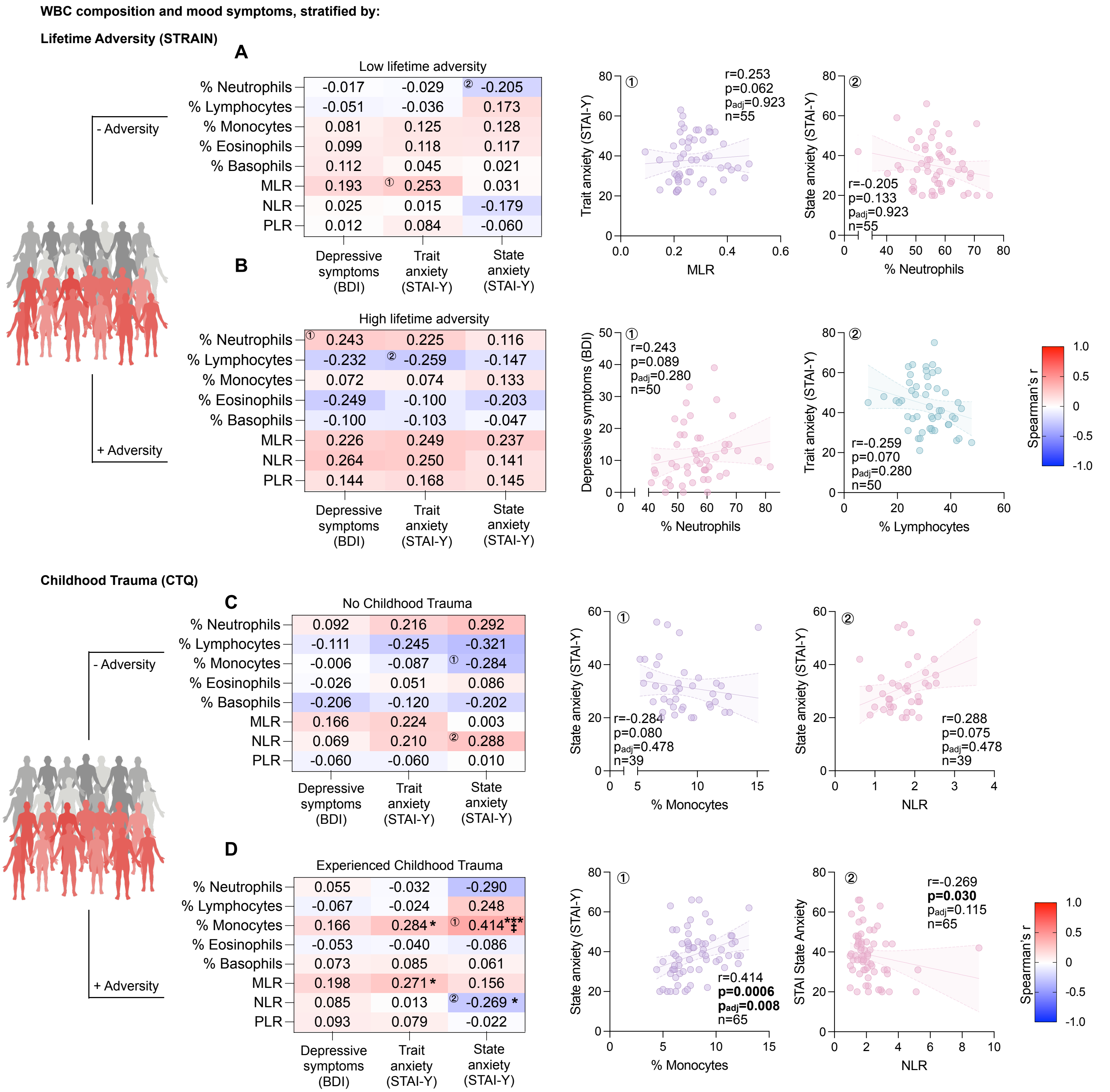


**Figure S2. Correlations between white blood cell composition with depression/anxiety, stratified by adversity exposure**

(A) Heatmap and select scatterplots of correlations between white blood cell population (WBC) percentages and monocyte lymphocyte ratio (MLR), neutrophil lymphocyte ratio (NLR), platelet lymphocyte ratio (PLR) with measures of depression and anxiety symptoms BDI, STAI- Trait, STAI -State) in MiSBIE participants who experienced low adversity (STRAIN severity score ≤44). n=51-55.

(B) Heatmap and select scatterplots of correlations between WBC percentages and ratios with measures of depression and anxiety symptoms (BDI, STAI- Trait, STAI -State) in MiSBIE participants who have experienced high adversity (STRAIN severity score >44). n=41-50.

(C) Heatmap and select scatterplots of correlations between white blood cell population (WBC) percentages and monocyte lymphocyte ratio (MLR), neutrophil lymphocyte ratio (NLR), platelet lymphocyte ratio (PLR) with measures of depression and anxiety symptoms (BDI, STAI- Trait, STAI -State) in MiSBIE participants who did not experience childhood trauma. n=39

(D) Heatmap and select scatterplots of correlations between WBC percentages and ratios with measures of depression and anxiety symptoms (BDI, STAI- Trait, STAI -State) in MiSBIE participants who experienced childhood trauma. n=65.

In scatterplots, linear regression lines shown for illustrative purposes only, all correlations are Spearman rank correlations. Effect size and p-values from Spearman rank correlation. *p<0.05, **p<0.01, ***p<0.001, ****p<0.0001. Benjamini-Hochberg FDR-corrected †$p_{adj}$<0.05, ‡$p_{adj}$<0.01, ††$p_{adj}$<0.001, ‡‡$p_{adj}$<0.0001.

#### Figure S3

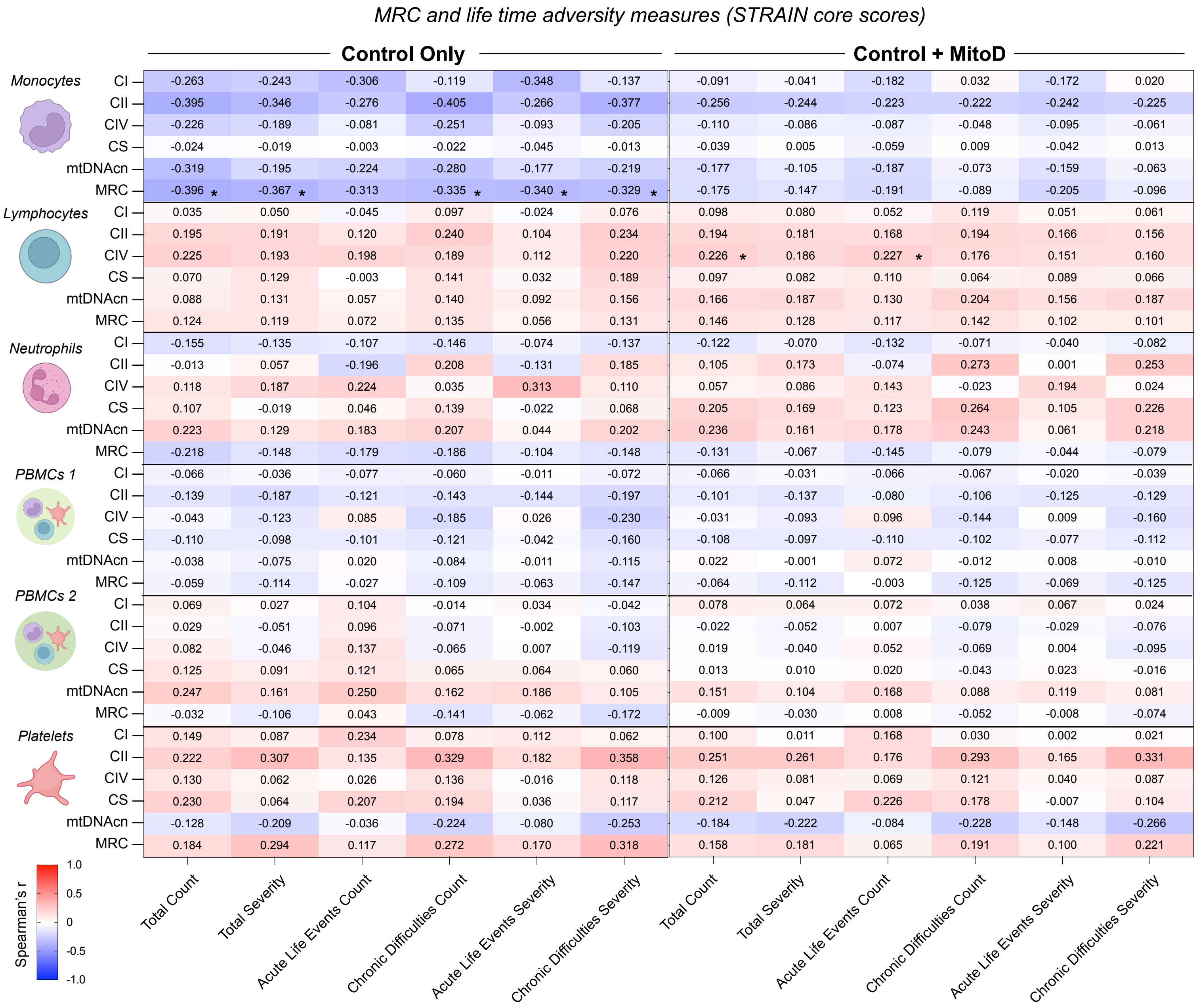


**Figure S3. Correlations between Mitochondrial variables of different cell types with psychosocial measures**
Correlations between CI, CII, CIV and CS enzyme activities, mtDNA and MRC of monocytes, lymphocytes, neutrophils, PBMCs (1/2), platelets and STRAIN Core subscales in the entire MiSBIE cohort (n=44-90) and in Control only (n=31-66).
Effect size and p-values from Spearman rank correlation. *p<0.05, **p<0.01, ***p<0.001, ****p<0.0001. No correlations remained significant following Benjamini-Hochberg FDR correction.

**Figure S4**

*MRC and childhood adversity measures (CTQ scores)*

**Control Only**

| Cell type | Variable | Physical Abuse | Emotional Abuse | Sexual Abuse | Emotional Neglect | Physical Neglect | Minimization Denial | CTQ Sum |
|---|---|---|---|---|---|---|---|---|
| *Monocytes* | CI | 0.197 | 0.086 | 0.019 | 0.156 | 0.192 | -0.218 | 0.184 |
| | CII | -0.235 | -0.440** | -0.098 | -0.195 | -0.301 | -0.017 | -0.330 |
| | CIV | -0.030 | -0.232 | 0.037 | -0.080 | -0.161 | -0.238 | -0.156 |
| | CS | -0.082 | 0.025 | 0.381 | 0.223 | 0.267 | 0.158 | 0.242 |
| | mtDNAcn | -0.173 | -0.184 | 0.034 | 0.049 | -0.001 | -0.310 | -0.071 |
| | MRC | 0.034 | -0.229 | -0.061 | -0.051 | -0.099 | -0.253 | -0.122 |
| *Lymphocytes* | CI | -0.018 | -0.045 | -0.116 | 0.026 | 0.131 | -0.106 | -0.028 |
| | CII | 0.003 | 0.051 | 0.036 | -0.014 | 0.082 | -0.048 | 0.004 |
| | CIV | 0.096 | -0.002 | 0.034 | -0.206 | -0.079 | -0.122 | -0.148 |
| | CS | 0.001 | -0.041 | -0.012 | -0.142 | -0.087 | -0.114 | -0.072 |
| | mtDNAcn | -0.002 | 0.055 | 0.098 | -0.027 | 0.064 | -0.037 | 0.033 |
| | MRC | -0.001 | -0.084 | -0.108 | -0.140 | -0.006 | 0.020 | -0.156 |
| *Neutrophils* | CI | 0.012 | -0.195 | 0.090 | -0.065 | 0.060 | 0.212 | -0.049 |
| | CII | 0.089 | 0.148 | 0.100 | 0.251 | 0.278 | -0.343 | 0.115 |
| | CIV | -0.125 | -0.012 | 0.133 | -0.026 | -0.016 | 0.306 | -0.014 |
| | CS | 0.043 | 0.230 | 0.230 | 0.207 | -0.003 | -0.043 | 0.145 |
| | mtDNAcn | -0.103 | 0.252 | 0.145 | 0.193 | -0.081 | -0.107 | 0.148 |
| | MRC | -0.040 | -0.247 | 0.024 | -0.122 | 0.050 | 0.169 | -0.135 |
| *PBMCs 1* | CI | 0.066 | 0.043 | -0.019 | 0.157 | 0.069 | -0.075 | 0.126 |
| | CII | -0.086 | -0.057 | 0.020 | 0.096 | 0.034 | -0.015 | -0.017 |
| | CIV | -0.105 | -0.045 | 0.139 | 0.082 | -0.178 | 0.041 | -0.052 |
| | CS | 0.042 | -0.030 | -0.004 | 0.195 | 0.012 | 0.050 | 0.060 |
| | mtDNAcn | -0.009 | 0.114 | 0.283 | 0.182 | 0.010 | -0.020 | 0.163 |
| | MRC | -0.046 | -0.082 | -0.036 | -0.039 | -0.043 | 0.026 | -0.104 |
| *PBMCs 2* | CI | -0.016 | 0.070 | 0.266 * | 0.139 | -0.077 | 0.049 | 0.026 |
| | CII | -0.157 | -0.020 | 0.280 * | -0.062 | -0.334** | 0.030 | -0.162 |
| | CIV | -0.095 | -0.082 | 0.332 * | -0.109 | -0.344** | 0.294 | -0.194 |
| | CS | -0.052 | 0.086 | 0.332 * | -0.047 | -0.276 * | 0.085 | -0.081 |
| | mtDNAcn | -0.024 | 0.045 | 0.243 | 0.074 | -0.071 | 0.110 | 0.016 |
| | MRC | -0.098 | -0.084 | 0.237 | -0.020 | -0.264 * | 0.145 | -0.147 |
| *Platelets* | CI | 0.098 | -0.074 | -0.014 | 0.227 | 0.104 | 0.273 | 0.142 |
| | CII | 0.010 | 0.002 | 0.010 | 0.042 | 0.066 | -0.038 | 0.186 |
| | CIV | -0.106 | -0.092 | -0.068 | -0.078 | 0.050 | 0.094 | 0.005 |
| | CS | -0.125 | -0.077 | 0.043 | 0.071 | 0.111 | 0.244 | 0.071 |
| | mtDNAcn | -0.083 | -0.166 | -0.116 | 0.033 | -0.110 | 0.240 | -0.009 |
| | MRC | 0.110 | 0.087 | 0.033 | 0.059 | 0.123 | -0.039 | 0.164 |

**Control + MitoD**

| Cell type | Variable | Physical Abuse | Emotional Abuse | Sexual Abuse | Emotional Neglect | Physical Neglect | Minimization Denial | CTQ Sum |
|---|---|---|---|---|---|---|---|---|
| *Monocytes* | CI | 0.284 * | 0.120 | -0.014 | 0.184 | 0.187 | -0.341 * | 0.237 |
| | CII | -0.208 | -0.385 ** | -0.018 | -0.144 | -0.237 | -0.008 | -0.247 |
| | CIV | -0.056 | -0.174 | 0.045 | -0.132 | -0.190 | -0.138 | -0.129 |
| | CS | 0.028 | -0.020 | 0.266 | 0.041 | 0.154 | 0.182 | 0.045 |
| | mtDNAcn | -0.108 | -0.138 | 0.103 | 0.044 | -0.026 | -0.276 * | -0.045 |
| | MRC | 0.068 | -0.143 | -0.084 | 0.009 | -0.066 | -0.326 * | -0.003 |
| *Lymphocytes* | CI | -0.005 | 0.003 | 0.070 | 0.051 | 0.134 | -0.095 | 0.060 |
| | CII | -0.032 | 0.010 | 0.129 | -0.037 | 0.045 | -0.059 | -0.005 |
| | CIV | 0.144 | 0.046 | 0.093 | -0.111 | 0.034 | -0.121 | -0.013 |
| | CS | -0.058 | -0.075 | 0.067 | -0.077 | -0.017 | -0.104 | -0.014 |
| | mtDNAcn | -0.033 | 0.095 | 0.135 | 0.036 | 0.073 | -0.094 | 0.079 |
| | MRC | 0.036 | -0.031 | 0.068 | -0.085 | 0.038 | -0.020 | -0.048 |
| *Neutrophils* | CI | 0.024 | -0.142 | 0.139 | -0.066 | 0.016 | 0.104 | -0.007 |
| | CII | -0.067 | 0.080 | 0.127 | 0.162 | 0.117 | -0.277 | 0.099 |
| | CIV | -0.111 | -0.012 | 0.261 | -0.098 | -0.100 | 0.217 | -0.048 |
| | CS | 0.084 | 0.202 | 0.274 * | 0.114 | -0.004 | -0.048 | 0.129 |
| | mtDNAcn | -0.071 | 0.145 | 0.121 | 0.099 | -0.073 | -0.144 | 0.086 |
| | MRC | -0.045 | -0.142 | 0.163 | -0.072 | 0.039 | 0.061 | -0.034 |
| *PBMCs 1* | CI | -0.013 | -0.055 | 0.025 | 0.044 | -0.043 | -0.015 | -0.013 |
| | CII | -0.138 | -0.086 | -0.018 | 0.060 | -0.028 | 0.006 | -0.083 |
| | CIV | -0.040 | -0.035 | -0.049 | 0.080 | -0.105 | 0.020 | -0.060 |
| | CS | -0.026 | -0.081 | 0.001 | 0.085 | -0.127 | 0.063 | -0.059 |
| | mtDNAcn | 0.089 | 0.132 | 0.087 | 0.203 | 0.053 | -0.028 | 0.167 |
| | MRC | -0.120 | -0.150 | -0.016 | -0.065 | -0.083 | 0.073 | -0.167 |
| *PBMCs 2* | CI | 0.047 | 0.059 | 0.180 | 0.137 | -0.040 | -0.053 | 0.022 |
| | CII | -0.172 | -0.140 | 0.120 | -0.120 | -0.356*** | 0.056 | -0.261 * |
| | CIV | -0.067 | -0.158 | 0.136 | -0.149 | -0.324** | 0.216 | -0.247 |
| | CS | -0.082 | -0.076 | 0.142 | -0.133 | -0.325** | 0.127 | -0.202 |
| | mtDNAcn | -0.042 | -0.090 | 0.077 | -0.011 | -0.146 | 0.143 | -0.106 |
| | MRC | -0.044 | -0.084 | 0.151 | -0.025 | -0.190 | 0.034 | -0.137 |
| *Platelets* | CI | 0.000 | -0.045 | -0.034 | 0.165 | 0.129 | 0.187 | 0.109 |
| | CII | -0.048 | -0.001 | 0.080 | 0.068 | -0.016 | -0.020 | 0.137 |
| | CIV | -0.149 | -0.030 | 0.044 | -0.163 | -0.037 | 0.037 | -0.026 |
| | CS | 0.035 | 0.002 | -0.076 | 0.136 | 0.223 | 0.107 | 0.139 |
| | mtDNAcn | -0.005 | -0.105 | -0.206 | 0.028 | 0.041 | 0.050 | -0.012 |
| | MRC | -0.009 | 0.029 | 0.170 | -0.023 | -0.039 | 0.015 | 0.086 |

Spearman's r (color scale): 1.0, 0.5, 0, -0.5, -1.0

**Figure S4. Correlations between Mitochondrial variables of different cell types with psychosocial measures**

Correlations between CI, CII, CIV and CS enzyme activities, mtDNA and MRC of monocytes, lymphocytes, neutrophils, PBMCs (1/2), platelets and CTQ sub scales in the entire MiSBIE cohort (n=44-90) and in Control only (n=30-65). Effect size and p-values from Spearman rank correlation. *p<0.05, **p<0.01, ***p<0.001, ****p<0.0001. No correlations remained significant following Benjamini-Hochberg FDR correction.

#### Figure S5

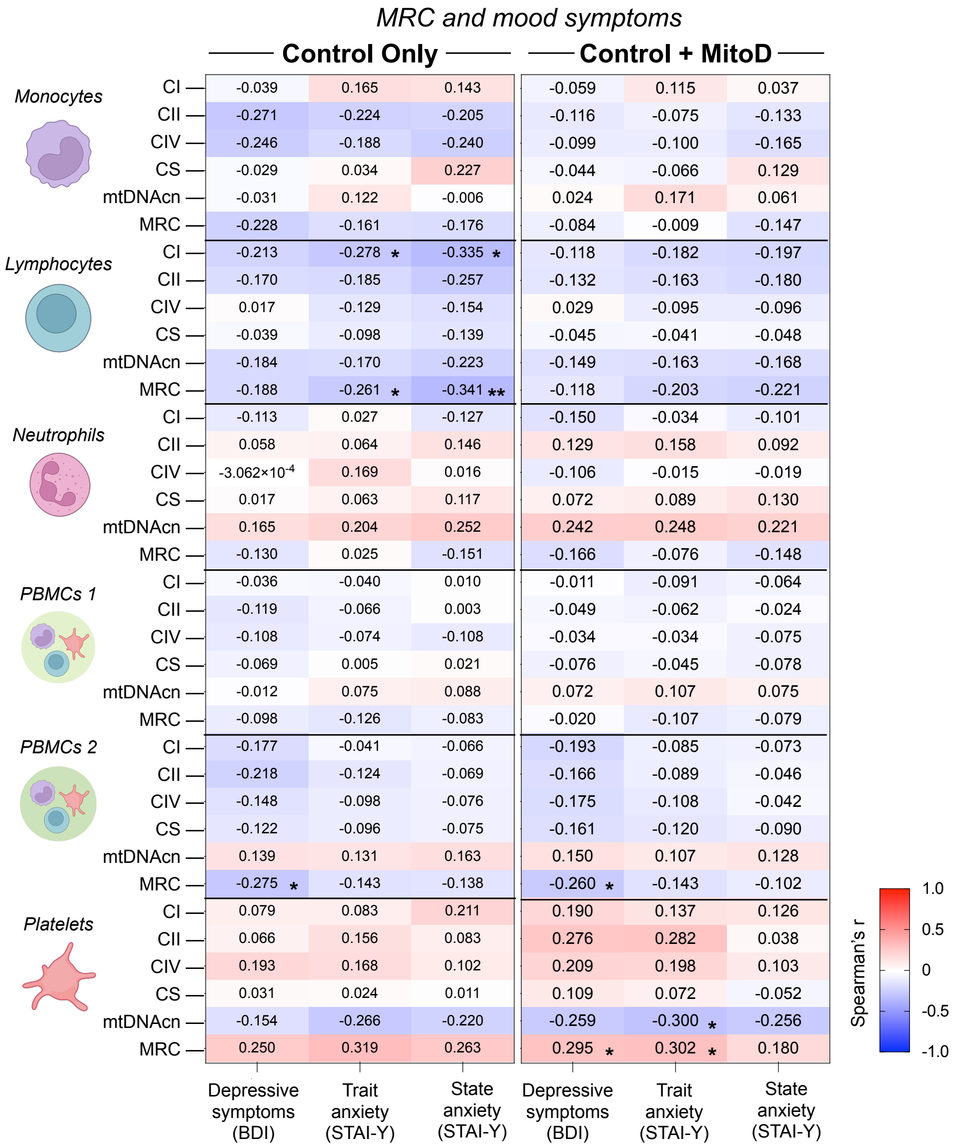


**Figure S5. Correlations between Mitochondrial variables of different cell types with psychosocial measures**

Correlations between CI, CII, CIV and CS enzyme activities, mtDNA and MRC of monocytes, lymphocytes, neutrophils, PBMCs (1/2), platelets and measures of depression and anxiety symptoms (BDI, STAI- Trait, STAI-State) in the entire MiSBIE cohort (n=44-91) and in Control only (n=30-65). Effect size and p-values from Spearman rank correlation. *p<0.05, **p<0.01, ***p<0.001, ****p<0.0001. No correlations remained significant following Benjamini-Hochberg FDR correction.

#### Figure S6

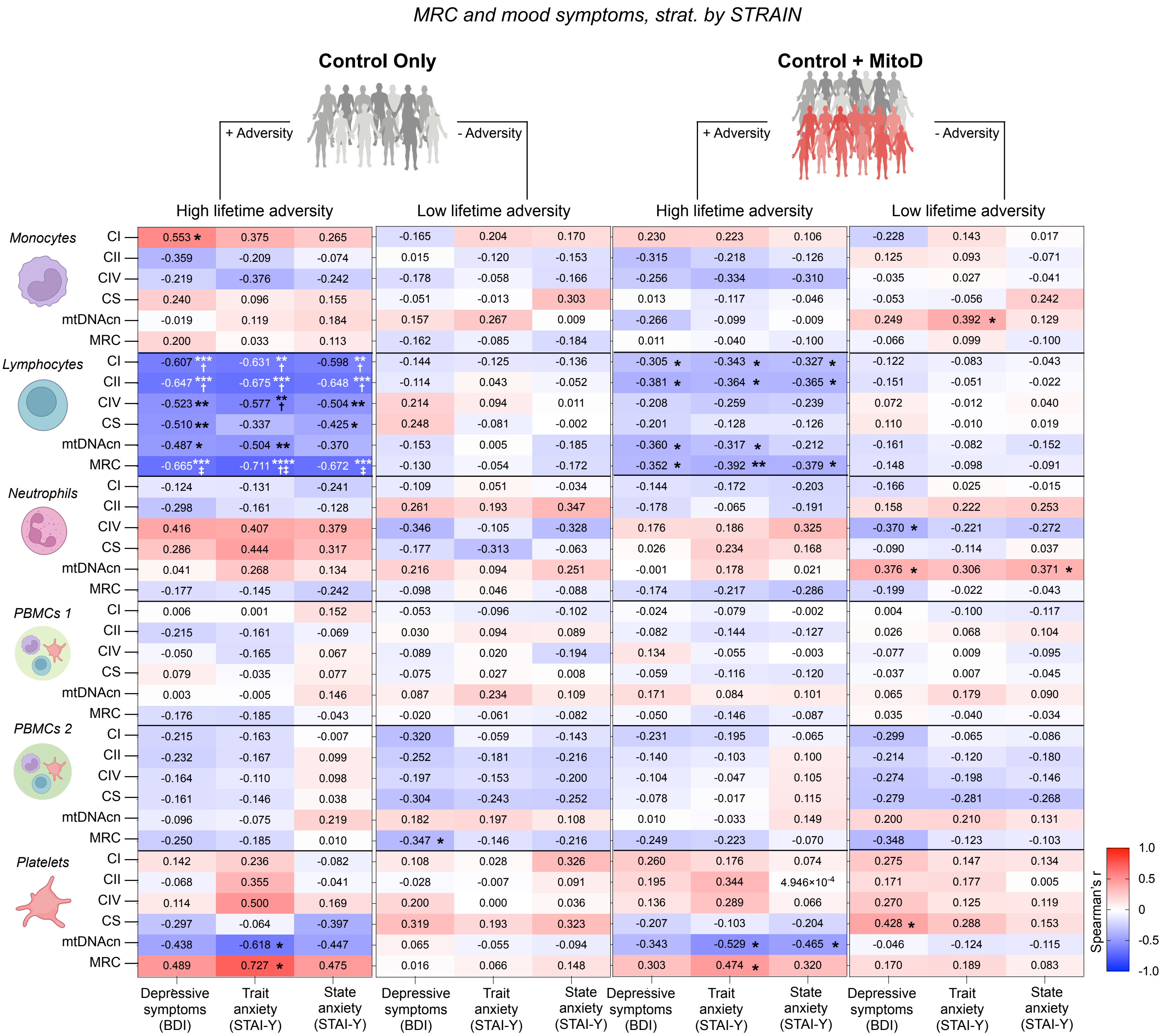


**Figure S6. Correlations between mitochondrial variables of different cell types with depression/anxiety, stratified Lifetime Adversity (STRAIN)**
Correlations between CI, CII, CIV and CS enzyme activities, mtDNA and MRC of monocytes, lymphocytes, neutrophils, PBMCs (1/2), platelets and measures of depression and anxiety symptoms (BDI, STAI- Trait, STAI-State) in MiSBIE participants with high adversity (STRAIN severity score >44) and low adversity (STRAIN severity score ≤44) in the entire MiSBIE cohort (low adversity n=24-48, high adversity n=20-44) and in control only (low adversity n=17-38, high adversity n=11-27). Effect size and p-values from Spearman rank correlation. *p<0.05, **p<0.01, ***p<0.001, ****p<0.0001. Benjamini-Hochberg FDR-corrected †$p_{adj}$<0.05, ‡$p_{adj}$<0.01, ††$p_{adj}$<0.001, ‡‡$p_{adj}$<0.0001.

**Figure S7**

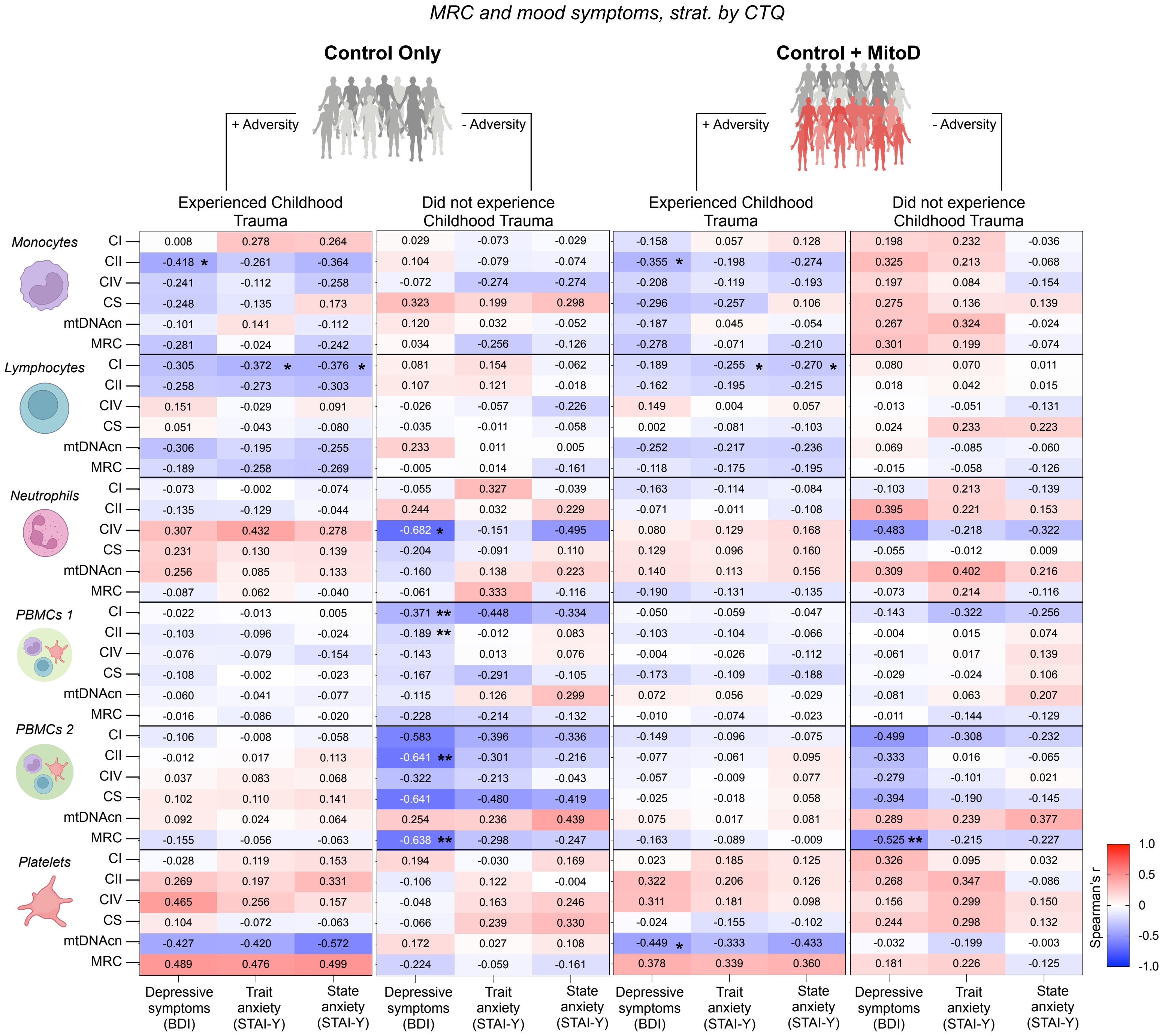


**Figure S7. Correlations between mitochondrial variables of different cell types with depression/anxiety, stratified by Childhood Trauma (CTQ)**
Correlations between CI, CII, CIV and CS enzyme activities, mtDNA and MRC of monocytes, lymphocytes, neutrophils, PBMCs (1/2), platelets and measures of depression and anxiety (BDI, STAI- Trait, STAI-State) in MiSBIE participants that did and did not experience childhood adversity in the entire MiSBIE cohort (experienced childhood trauma n=23-57, did not experience childhood trauma n=21-33) and in controls only (experienced childhood trauma n=19-43, did not experience childhood trauma n=11-22). (defined cutoffs from Nakajima 2022). Effect size and p-values from Spearman rank correlation. *p<0.05, **p<0.01, ***p<0.001, ****p<0.0001.No correlations remained significant following Benjamini-Hochberg FDR correction.

**Figure S8**

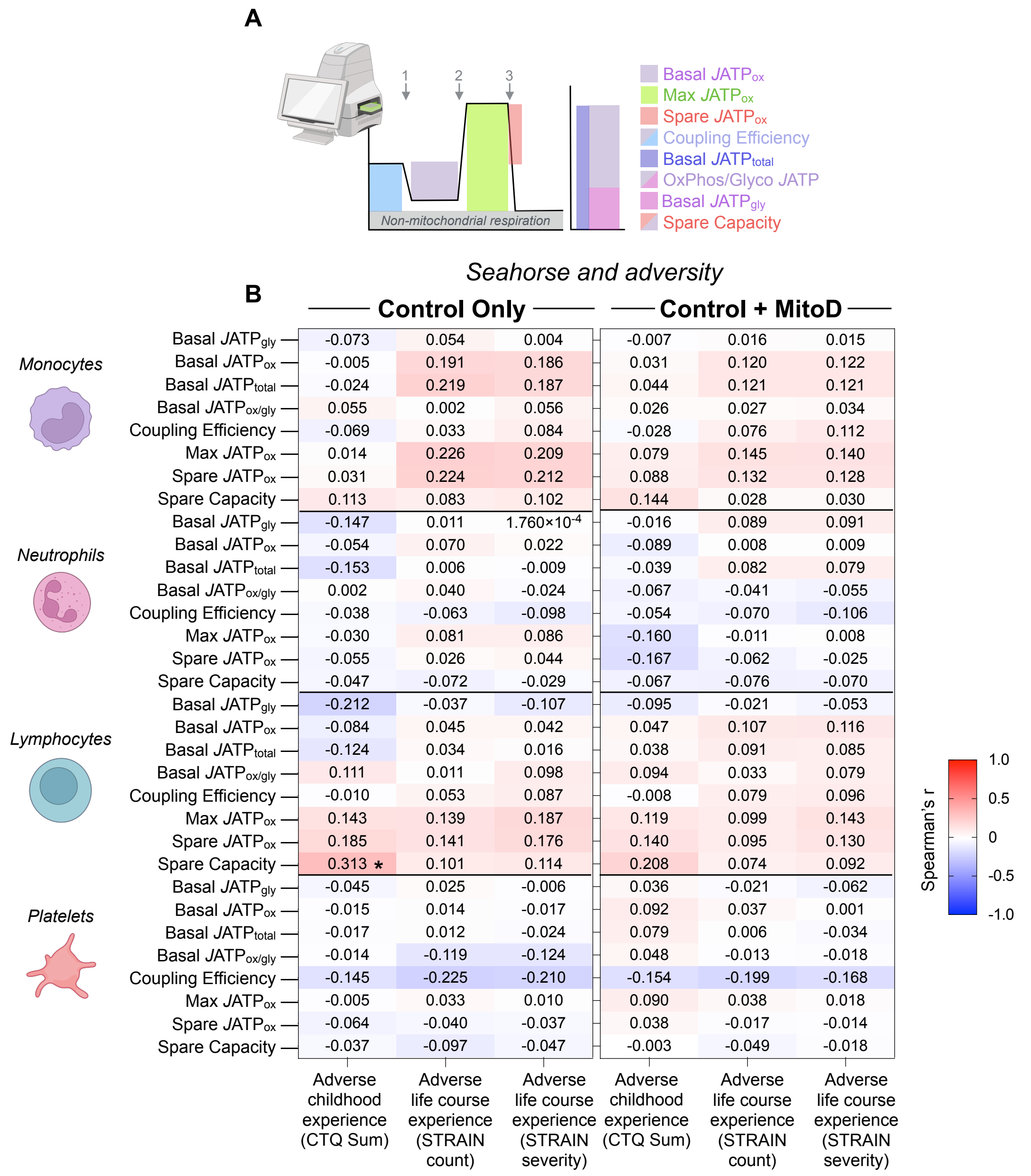


**Figure S8. Correlations between respirometry variables and adversity measures**

(A) Schematic of respirometry trace (left), completed in the Seahorse XF Flux analyzer, measuring oxygen consumption with the addition of 1. Oligomycin, 2. FCCP, 3. Rotenone and Antimycin A. Schematic of total ATP production (right), consisting of ATP produced via OxPhos and glycolysis.

(B) Heatmap of correlations between ATP production rates derived from mitochondrial OxPhos under both basal (Basal JATPox), ATP production rates derived from cytosolic glycolysis under basal conditions (basal JATPgly), total ATP cellular ATP production rate (basal JATPtotal), the ratio of basal ATP production rate derived from mitochondrial OxPhos over basal ATP production rate derived from cytosolic glycolysis (Basal JATPox/gly), the percentage of basal oxygen consumption linked to ATP production (Coupling Efficiency), ATP production rate derived from maximal (Max JATPox) respiration, the ATP production rate derived from OxPhos under maximal respiratory activity without accounting for basal ATP production derived from OxPhos (Spare JATPox), and the Spare JATPox expressed as a percentage of basal Basal JATPox (Spare JATPox Capacity) with measures of adversity (CTQ, STRAIN Total Count, STRAIN Total Severity) in controls only (n=60-66), and in the entire MiSBIE cohort (n=92-95). Effect size and p-values from Spearman rank correlation. *p<0.05, **p<0.01, ***p<0.001, ****p<0.0001. No correlations remained significant following Benjamini-Hochberg FDR correction.

#### Figure S9

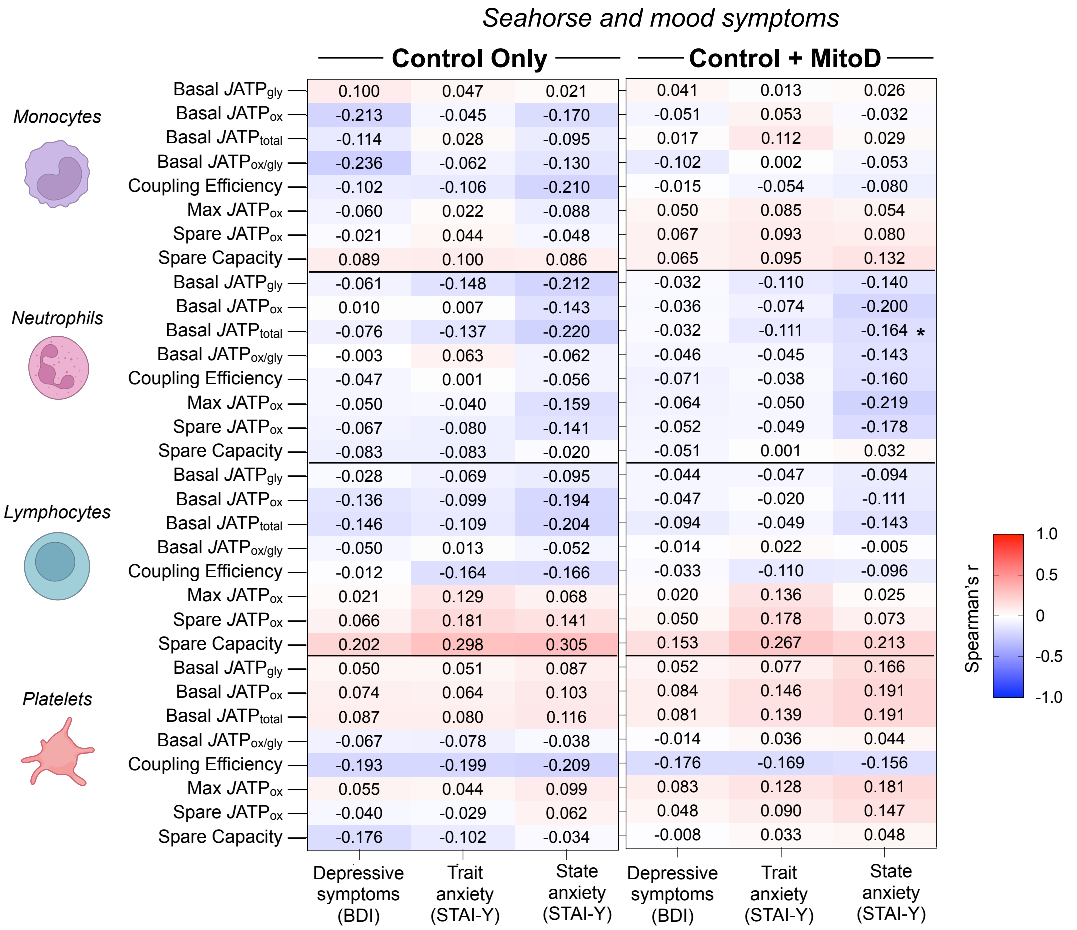


**Figure S9. Correlations between respirometry variables and mood symptoms**

Heatmap of correlations between ATP production rates derived from mitochondrial OxPhos under both basal (Basal JATPox), ATP production rates derived from cytosolic glycolysis under basal conditions (basal JATPgly), total ATP cellular ATP production rate (basal JATPtotal), the ratio of basal ATP production rate derived from mitochondrial OxPhos over basal ATP production rate derived from cytosolic glycolysis (Basal JATPox/gly), the percentage of basal oxygen consumption linked to ATP production (Coupling Efficiency), ATP production rate derived from maximal (Max JATPox) respiration, the ATP production rate derived from OxPhos under maximal respiratory activity without accounting for basal ATP production derived from OxPhos (Spare JATPox), and the Spare JATPox expressed as a percentage of basal Basal JATPox (Spare JATPox Capacity) with measures of depression and anxiety (BDI, STAI-Trait, STAI-State) in controls only (n=61-65), and in the entire MiSBIE cohort (n=92-97). Effect size and p-values from Spearman rank correlation. *p<0.05, **p<0.01, ***p<0.001, ****p<0.0001. No correlations remained significant following Benjamini-Hochberg FDR correction.

**Figure S10**

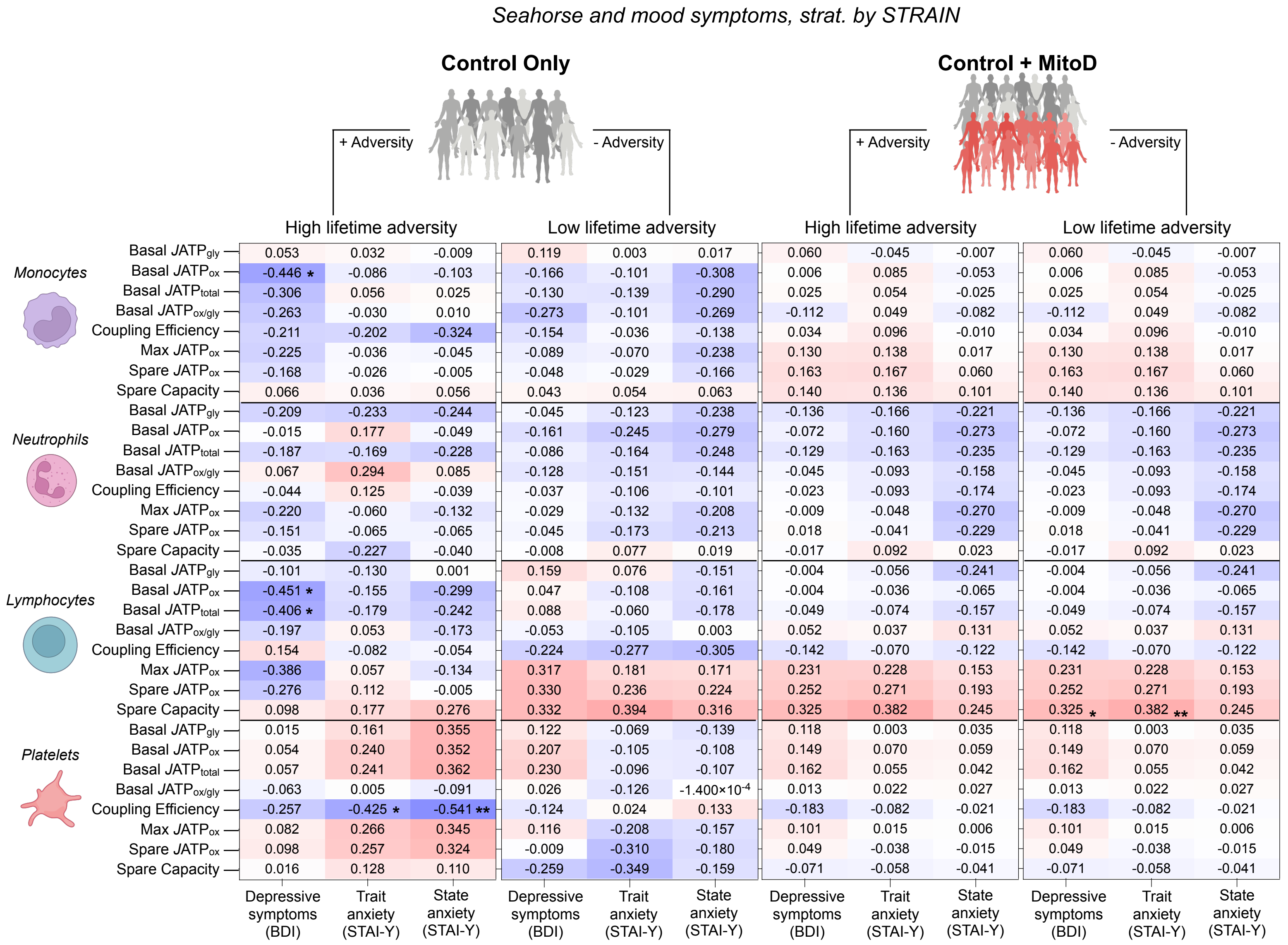


**Figure S10. Correlations between respirometry variables and mood symptoms, stratified by Lifetime Adversity (STRAIN)**
Heatmap of correlations between ATP production rates derived from mitochondrial OxPhos under both basal (Basal JATPox), ATP production rates derived from cytosolic glycolysis under basal conditions (basal JATPgly), total ATP cellular ATP production rate (basal JATPtotal), the ratio of basal ATP production rate derived from mitochondrial OxPhos over basal ATP production rate derived from cytosolic glycolysis (Basal JATPox/gly), the percentage of basal oxygen consumption linked to ATP production (Coupling Efficiency), ATP production rate derived from maximal (Max JATPox) respiration, the ATP production rate derived from OxPhos under maximal respiratory activity without accounting for basal ATP production derived from OxPhos (Spare JATPox), and the Spare JATPox expressed as a percentage of basal Basal JATPox (Spare JATPox Capacity) with measures of depression and anxiety (BDI, STAI-Trait, STAI-State) in MiSBIE participants with high adversity (STRAIN severity score >44) and low adversity (STRAIN severity score ≤44) in the entire MiSBIE cohort (low adversity n=47–50, high adversity n=44-47) and in control only (low adversity n=34-36, high adversity n=27-29). Effect size and p-values from Spearman rank correlation. *p<0.05, **p<0.01, ***p<0.001, ****p<0.0001. No correlations remained significant following Benjamini-Hochberg FDR correction.

#### Figure S11

*Seahorse and mood symptoms, strat. by CTQ*

| | | **Control Only** | | | | | | **Control + MitoD** | | | | | |
|---|---|---|---|---|---|---|---|---|---|---|---|---|---|
| | | + Adversity: Experienced Childhood Trauma | | | - Adversity: Did not experience Childhood Trauma | | | + Adversity: Experienced Childhood Trauma | | | - Adversity: Did not experience Childhood Trauma | | |
| Cell type | Variable | Depressive symptoms (BDI) | Trait anxiety (STAI-Y) | State anxiety (STAI-Y) | Depressive symptoms (BDI) | Trait anxiety (STAI-Y) | State anxiety (STAI-Y) | Depressive symptoms (BDI) | Trait anxiety (STAI-Y) | State anxiety (STAI-Y) | Depressive symptoms (BDI) | Trait anxiety (STAI-Y) | State anxiety (STAI-Y) |
| *Monocytes* | Basal $JATP_{gly}$ | 0.117 | 0.046 | 0.003 | 0.156 | 0.098 | 0.172 | 0.061 | -0.068 | -0.104 | -0.032 | 0.098 | 0.177 |
| | Basal $JATP_{ox}$ | -0.126 | -0.034 | -0.145 | -0.323 | -0.076 | -0.216 | 0.024 | 0.069 | -0.021 | -0.178 | -0.043 | -0.133 |
| | Basal $JATP_{total}$ | -0.024 | 0.062 | -0.099 | -0.180 | -0.001 | -0.039 | 0.106 | 0.088 | -0.047 | -0.170 | 0.049 | 0.029 |
| | Basal $JATP_{ox/gly}$ | -0.223 | -0.069 | -0.108 | -0.285 | -0.130 | -0.286 | -0.091 | 0.068 | 0.062 | -0.025 | -0.108 | -0.242 |
| | Coupling Efficiency | -0.033 | 0.017 | -0.167 | -0.338 | -0.278 | -0.268 | 0.059 | 0.036 | -0.060 | -0.157 | -0.172 | -0.073 |
| | Max $JATP_{ox}$ | 0.045 | 0.090 | -0.068 | -0.191 | -0.016 | -0.147 | 0.166 | 0.151 | 0.095 | -0.112 | 0.054 | -0.008 |
| | Spare $JATP_{ox}$ | 0.080 | 0.119 | -0.022 | -0.170 | 0.002 | -0.127 | 0.174 | 0.153 | 0.119 | -0.105 | 0.046 | -0.003 |
| | Spare Capacity | 0.102 | 0.136 | 0.080 | 0.000 | 0.025 | -0.025 | 0.067 | 0.099 | 0.137 | -0.008 | 0.124 | 0.109 |
| *Neutrophils* | Basal $JATP_{gly}$ | -0.148 | -0.348 * | -0.307 * | 0.165 | 0.393 | 0.004 | -0.052 | -0.262 * | -0.259 * | -0.063 | 0.135 | -0.062 |
| | Basal $JATP_{ox}$ | 0.135 | 0.058 | -0.174 | -0.216 | 0.076 | 0.068 | 0.010 | -0.023 | -0.161 | -0.141 | -0.170 | -0.230 |
| | Basal $JATP_{total}$ | -0.126 | -0.320 * | -0.336 * | 0.032 | 0.368 | 0.022 | -0.027 | -0.234 | -0.267 | -0.090 | 0.107 | -0.094 |
| | Basal $JATP_{ox/gly}$ | 0.145 | 0.173 | -0.028 | -0.216 | 0.040 | 0.091 | 0.006 | 0.034 | -0.065 | -0.088 | -0.118 | -0.142 |
| | Coupling Efficiency | -0.047 | -0.060 | -0.162 | -0.009 | 0.281 | 0.268 | -0.075 | -0.083 | -0.215 | -0.038 | 0.135 | 0.059 |
| | Max $JATP_{ox}$ | 0.060 | 0.041 | -0.123 | -0.378 | -0.228 | -0.246 | -0.043 | -0.037 | -0.156 | -0.154 | -0.025 | -0.274 |
| | Spare $JATP_{ox}$ | 0.025 | 0.005 | -0.093 | -0.447 | -0.392 | -0.365 | -0.019 | -0.037 | -0.108 | -0.192 | -0.045 | -0.301 |
| | Spare Capacity | -0.085 | 0.001 | 0.031 | -0.080 | -0.386 | -0.246 | -0.058 | 0.012 | 0.045 | -0.034 | 0.040 | 0.024 |
| *Lymphocytes* | Basal $JATP_{gly}$ | -0.104 | -0.119 | -0.102 | 0.419 | 0.263 | 0.088 | -0.108 | -0.116 | -0.140 | 0.124 | 0.084 | -0.121 |
| | Basal $JATP_{ox}$ | -0.033 | 0.021 | -0.083 | -0.310 | -0.157 | -0.370 | 0.021 | 0.068 | -0.026 | -0.244 | -0.228 | -0.399 |
| | Basal $JATP_{total}$ | -0.074 | -0.042 | -0.128 | -0.173 | -0.052 | -0.296 | -0.062 | -0.020 | -0.097 | -0.192 | -0.175 | -0.377 |
| | Basal $JATP_{ox/gly}$ | 0.043 | 0.122 | 0.018 | -0.465 * | -0.273 | -0.266 | 0.068 | 0.139 | 0.088 | -0.229 | -0.197 | -0.149 |
| | Coupling Efficiency | 0.171 | 0.041 | 0.042 | -0.341 | -0.370 | -0.284 | 0.079 | 0.001 | 0.036 | -0.194 | -0.208 | -0.164 |
| | Max $JATP_{ox}$ | 0.029 | 0.099 | 0.078 | -0.148 | 0.038 | -0.188 | -0.024 | 0.071 | 0.057 | -0.056 | 0.103 | -0.200 |
| | Spare $JATP_{ox}$ | 0.055 | 0.135 | 0.120 | -0.071 | 0.076 | -0.090 | -0.022 | 0.093 | 0.083 | 0.014 | 0.161 | -0.127 |
| | Spare Capacity | 0.031 | 0.072 | 0.123 | 0.226 | 0.283 | 0.242 | -0.068 | -0.009 | 0.042 | 0.282 | 0.460 | 0.288 |
| *Platelets* | Basal $JATP_{gly}$ | 0.026 | -0.041 | 0.007 | 0.083 | 0.206 | 0.293 | 0.099 | 0.048 | 0.123 | -0.036 | 0.201 | 0.331 |
| | Basal $JATP_{ox}$ | 0.193 | 0.121 | 0.201 | -0.221 | -0.059 | -0.081 | 0.185 | 0.148 | 0.197 | -0.095 | 0.119 | 0.135 |
| | Basal $JATP_{total}$ | 0.184 | 0.105 | 0.181 | -0.114 | 0.043 | 0.025 | 0.179 | 0.139 | 0.188 | -0.111 | 0.105 | 0.140 |
| | Basal $JATP_{ox/gly}$ | 0.082 | 0.075 | 0.183 | -0.348 | -0.315 | -0.409 | 0.085 | 0.110 | 0.129 | -0.149 | -0.073 | -0.112 |
| | Coupling Efficiency | -0.213 | -0.156 | -0.172 | -0.048 | -0.159 | -0.177 | -0.115 | -0.096 | -0.085 | -0.162 | -0.159 | -0.168 |
| | Max $JATP_{ox}$ | 0.211 | 0.105 | 0.211 | -0.322 | -0.151 | -0.179 | 0.199 | 0.154 | 0.217 | -0.152 | 0.059 | 0.036 |
| | Spare $JATP_{ox}$ | 0.129 | 0.041 | 0.218 | -0.348 | -0.116 | -0.194 | 0.161 | 0.123 | 0.223 | -0.124 | 0.094 | 0.003 |
| | Spare Capacity | -0.083 | -0.057 | 0.089 | -0.307 | -0.110 | -0.287 | 0.038 | 0.075 | 0.159 | -0.017 | 0.078 | -0.187 |

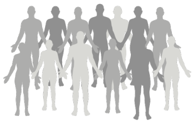

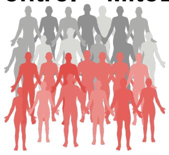

**Figure S11. Correlations between respirometry variables and mood symptoms, stratified by Childhood Adversity (CTQ)**
Heatmap of correlations between ATP production rates derived from mitochondrial OxPhos under both basal (Basal JATPox), ATP production rates derived from cytosolic glycolysis under basal conditions (basal JATPgly), total ATP cellular ATP production rate (basal JATPtotal), the ratio of basal ATP production rate derived from mitochondrial OxPhos over basal ATP production rate derived from cytosolic glycolysis (Basal JATPox/gly), the percentage of basal oxygen consumption linked to ATP production (Coupling Efficiency), ATP production rate derived from maximal (Max JATPox) respiration, the ATP production rate derived from OxPhos under maximal respiratory activity without accounting for basal ATP production derived from OxPhos (Spare JATPox), and the Spare JATPox expressed as a percentage of basal Basal JATPox (Spare JATPox Capacity) with measures of depression and anxiety (BDI, STAI-Trait, STAI-State) in n MiSBIE participants that did and did not experience childhood adversity in the entire MiSBIE cohort (experienced childhood trauma n=59-64, did not experience childhood trauma n=31-34) and in controls only (experienced childhood trauma n=39-44, did not experience childhood trauma n=19-21). (defined cutoffs from Nakajima 2022). Effect size and p-values from Spearman rank correlation. *p<0.05, **p<0.01, ***p<0.001, ****p<0.0001. No correlations remained significant following Benjamini-Hochberg FDR correction.
