## Supplemental Table 1 for "Lifetime adversity exposure, mood symptoms, and immune mitochondrial bioenergetics"

**Table S1. Participant characteristics.** Demographic, immune cell proportion, mood symptom and adversity data in healthy controls and individuals with mitochondrial disease (MitoD). Data shown as mean (SD), except for Sex and Race/Ethnicity, which is shown as count. Tests indicated below.

|  | Controls | MitoD | U* | p-value |
| --- | --- | --- | --- | --- |
| Number of Participants | 68 | 37 |  |  |
| Age | 37.28 (10.67) | 38.14 (10.96) | 1210 | 0.747 ^a^ |
| Sex |  |  |  | >0.99 ^b^ |
| Female | 46 | 25 |  |  |
| Male | 22 | 12 |  |  |
| BMI | 25.14 (5.31) | 23.73 (5.15) | 1072 | 0.213 ^a^ |
| Race/Ethnicity |  |  |  | 0.063 ^b^ |
| American Indian/Alaskan Native | 1 | 1 |  |  |
| Asian | 5 | 1 |  |  |
| Black/African American | 12 | 2 |  |  |
| Hispanic/Latinx | 8 | 1 |  |  |
| Native Hawaiian/Other Pacific Islander | 0 | 0 |  |  |
| White | 46 | 35 |  |  |
| Mood Symptoms |  |  |  |  |
| Depressive Symptoms (BDI) | 8.08 (8.38) | 10.4 (9.06) | 996.5 | 0.099 ^a^ |
| Trait Anxiety Symptoms (STAI-Y) | 40.7 (12.9) | 41.2 (11.1) | 1186 | 0.716 ^a^ |
| State Anxiety Symptoms (STAI-Y) | 36.7 (12.6) | 35.3 (9.84) | 1199 | 0.785 ^a^ |
| Adversity |  |  |  |  |
| STRAIN Total Stressor Count | 19.1 (13.3) | 20.5 (11.8) | 1075 | 0.423 ^a^ |
| STRAIN Total Stressor Severity | 48.3 (31.5) | 53.5 (30.3) | 1043 | 0.308 ^a^ |
| CTQ Sum Score | 37.8 (15.0) | 36.7 (13.5) | 1245 | 0.932 ^a^ |

* Mann-Whitney U for continuous variables only

^a^ Mann-Whitney test

^b^ Fisher’s exact test
